# Perceptual Differences between Cortical and Peripheral Stimulation Strategies for Sensory Restoration

**DOI:** 10.1101/2025.08.20.25334094

**Authors:** Brianna C. Hutchison, Rohit Bose, Bronwyn J. Spilker, Preethisiri Bhat, William D. Memberg, Aaron Ketting-Olivier, Eric Herring, Jennifer Sweet, Jonathan P. Miller, Robert F. Kirsch, A. Bolu Ajiboye, Emily L. Graczyk

## Abstract

Both intracortical microstimulation (ICMS) and peripheral nerve stimulation (PNS) can restore tactile sensation to people living with physical disabilities, such as spinal cord injury (SCI) or amputation. While both techniques have demonstrated success in evoking meaningful sensations in the upper limb, they have only been investigated in separate studies with different patient populations, and thus their perceptual characteristics have never been systematically compared to determine the relative advantages and limitations of each approach. In this study, we directly compared the perceived sensations evoked by ICMS and PNS to those evoked by mechanical touch in a participant with sensory incomplete spinal cord injury. We observed that ICMS evoked more localized percepts and felt more qualitatively similar to natural touch than PNS, whereas PNS evoked higher intensity and more reliable percepts than ICMS. We also found that, across stimulation approaches, the perceived naturalness ratings of sensations were more strongly related to the other perceptual variables than to stimulation variables, suggesting that naturalness is a higher order perceptual dimension that is cognitively constructed by the participant. Our results indicate that ICMS may be the better sensory stimulation approach for conveying naturalistic touch experiences during haptic exploration, due to its similarity to mechanical touch in perceptual quality. In contrast, PNS may be the better stimulation modality to deliver consistent and effective sensory feedback during closed-loop functional tasks, due to its higher reliability. These insights provide a framework for future development of patient-specific sensory neuroprostheses based on the needs and goals of the user.

## Introduction

Tactile feedback to the hand plays an essential role in activities of daily living^1^ by signaling object contact^2^, indicating the amount of force applied by the hand and fingers^3^, and providing detailed information about the physical properties of grasped objects^4^. Damage to the somatosensory pathway, such as spinal cord injury (SCI) or amputation, impairs function related to sensing and interacting with objects and can also lead to negative psychological effects such as increased feelings of isolation, depression, and anxiety^5,6^. Recent advances in implantable neural interfacing systems have shown that electrically stimulating various parts of the somatosensory pathway can restore the missing or impaired sensations^7^. The location of stimulation along the pathway typically depends on the location and nature of the neurological injury of the patient, as well as ethical concerns about the possibility of the implanted device damaging functional structures that would further impair the patient^8–10^. Specifically, systems to restore somatosensory feedback to people with limb loss tend to involve peripheral nerve stimulation (PNS) of nerves in the amputated limb^11–14^, while systems to restore somatosensation to people with SCI tend to involve intracortical microstimulation (ICMS) of primary somatosensory cortex (S1)^15–18^.

Many prior studies have demonstrated that the sensory percepts produced by both PNS and ICMS can be modulated along several perceptual dimensions, conveying useful information during grasp and haptic exploration^7,15,19–21^. For example, modulating the stimulation magnitude of ICMS^22^ and PNS^23–25^ has been shown to modify the perceived intensity of the evoked sensation. The temporal properties of a sensation can also be modulated by changing the frequency and pattern of stimulation pulses for both ICMS^26,27^ and PNS^28,29^. The evoked tactile percepts have also been shown to be functionally useful in grasp and manipulation tasks with a robotic or prosthetic hand. ICMS can be mapped to pressure sensors in real-time, improving the ability of SCI participants to grasp and transfer objects using robotic limbs^30^. People with limb loss using sensorized prostheses providing touch feedback via PNS were also able to modulate force applied by the prosthesis^11,24,31^, grasp delicate objects^11^, dexterously manipulate objects^25^, and discriminate objects with high accuracy^14,25,31,32^.

One of the major challenges in the field of somatosensory neuroprostheses is that neither PNS nor ICMS consistently evoke sensations that feel completely natural. For example, the projected fields evoked by either PNS or ICMS tend to be larger than the minimum percept size that can be detected with mechanical indentations in the intact sensory system^33,34^. In addition, the perceived qualities of both ICMS and PNS frequently include descriptors such as tingling^11,12,15,26,35^, paresthesia^35^, squeezing^20,36^, buzzing^12,26^, vibrating^12,15,26,37^, and electrical^12,15,23^, which are not typical descriptions of natural touch sensation^38^. In a prior study, ICMS could be reliably discriminated from mechanical vibration^18^, indicating ICMS does not perfectly replicate normal touch. PNS is also reported to feel unnatural using traditional stimulation patterns^11,39,40^, although naturalness appears to improve with more complex, “biomimetic” stimulation patterns for both PNS and ICMS^17,39,41^. The challenge of achieving natural sensation from neurostimulation is further complicated by the ambiguity of what “naturalness” means, given that it is not a primary dimension of sensation in classical psychophysics^42^. It is unclear whether perceived naturalness can be rated independently of the other primary perceptual dimensions (e.g. intensity, location, and quality) and whether the definition of naturalness is consistent across studies and participants.

While PNS has primarily been studied in people with limb loss, it could be beneficial for people with other types of injuries, such as incomplete SCI. Approximately 47% of people with tetraplegia retain at least partial sensory function below injury^11,43^ because they have an incomplete injury resulting in partially preserved sensory pathways. However, although portions of the sensory pathways remain intact, people with incomplete SCI typically have deficits in one or more sensory sub-modalities, including reduced sensitivity or acuity in touch, vibration, and/or temperature^44^. Thus, PNS could be a viable strategy to restore sensory feedback for people with incomplete SCI by augmenting existing neural activity^37^ and/or by serving as a supplemental information channel to augment the person’s residual sensory capabilities. In addition, surgeries to implant electrodes for PNS can be less complex and invasive than the craniotomy required for ICMS^36,45^. PNS for sensory restoration could be used in conjunction with less invasive motor restoration options, such as functional electrical stimulation (FES)^43^, surgical interventions, or other assistive devices, resulting in a bidirectional sensorimotor system that is less invasive than intracortical brain computer interfacing systems (BCI). In addition to evoking sensation by activating below-injury pathways, PNS may also have long-term benefits by promoting plasticity to improve residual sensory function^46–48^.

While prior studies indicate both ICMS and PNS are viable options for somatosensory restoration, they have yet to be directly compared in a single study. Thus, it remains unclear how their capabilities and outcomes truly compare to one another. Past studies have implemented PNS and ICMS in separate patient populations and use different methodologies for assessment of sensation. For example, people with unilateral amputations can typically move their other hand to circle the projected fields^11,35^, while people with motor-complete SCI cannot move their hands at all and often describe the projected fields by verbally reporting discretized regions^15,36^. Different studies also evaluate characteristics of sensation differently. For example, tactile quality information may be described by the participant using their own words^11,20^, selecting words from a study-specific descriptor word bank^12,15^, or numerically rating how applicable a descriptor word was to a stimulation-evoked sensation^38^. Additionally, quality word choice varies highly across participants, channels, and stimulation parameters^12,26^. Therefore, the comparisons between ICMS, PNS, and natural touch are obfuscated by the heterogeneity in the methods of measuring tactile percepts across studies, the variation in reported percepts across participants, and the idiosyncrasies of channel-specific and parameter-specific percepts. To understand how to better translate somatosensory neuroprostheses to the clinic, the sensations produced by ICMS and PNS need to be directly compared within the same participants and using the same methodology to better understand the perceptual capabilities of each stimulation approach.

In this study, we directly compare sensory percepts evoked by ICMS, PNS, and mechanical indentation of the hand in a participant with sensory-incomplete, motor-complete SCI (C3/C4 AIS-B SCI, age in 20s, male) enrolled in the Reconnecting the Hand and Arm to the Brain (ReHAB) clinical trial. As part of his implanted system, he received two 64-channel microelectrode arrays in Brodmann’s area 1 of S1 for ICMS and 16-channel Composite Flat Interface Nerve Electrodes (C-FINEs) for PNS around his median and ulnar nerves in the contralateral arm^36^(**Supplementary Figure 1).** Since the ReHAB participant has a sensory incomplete SCI, he can feel mechanical indentation of his hand as well as sensory percepts elicited by PNS applied to the nerves innervating his hand^36^. The direct comparison between ICMS, PNS, and mechanical touch in the same person is a novel opportunity to explore the relative benefits of these stimulation modalities for restoring the sense of touch to people with incomplete SCI. Findings from this cross-modality study can inform sensory neuroprosthesis selection for other people with sensory-incomplete SCIs and could be extended to other patient populations experiencing sensory loss from amputation, stroke, or other neuropathies.

## Results

A male participant with a sensory-incomplete high-cervical spinal cord injury (C3/C4 AIS-B) compared tactile sensations produced by ICMS, PNS, and a computer-actuated mechanical indenter (**Figure 1**). The participant retained partial sensation in his hands post-SCI, as evidenced by the results of the monofilament detection test and the two-point discrimination test both before and after his implantation surgery (**Supplementary Table 1**). For ICMS, 39% and 23% of contacts in the medial and lateral array, respectively, evoked sensation on the hand^36^. For PNS, 40% and 47% of C-FINE contacts on the median and ulnar nerve, respectively, evoked sensation on the hand. Among those contacts that evoked consistent sensations, two electrodes in the S1 intracortical arrays - one on the lateral array (ICMS_LE10) and one on the medial array (ICMS_ME59) - and two C-FINE contacts - one on the median nerve (PNS_MC5) and one on the ulnar nerve (PNS_UC4) - were selected for this study. C-FINE contacts were selected to best approximate the positions of the sensations evoked by ICMS, though no C-FINE contacts produced percepts that exactly overlapped the ICMS percept locations. A mechanical indenter was positioned so that it delivered mechanical indentation (Mech) stimuli to regions of the participant’s palm or fingers that overlapped with the ICMS-evoked percepts (Mech_ICMS_LE10_ and Mech_ICMS_ME59_) or PNS-evoked percepts (Mech_PNS_MC5_ and Mech_PNS_UC4_) (**Figure 2A,B**). The dynamic range was determined for each contact and mechanical indenter position, defined as the range from detection threshold to the participant’s self-selected maximum comfortable level. Then, three suprathreshold stimulation levels were selected for each modality: 30%, 60%, and 90% of the dynamic range (referred to as low, medium and high). In each trial, stimulation was applied via a single modality at a time. Stimulation modalities and levels were delivered in a randomized order, and the participant reported the stimulation modality, perceived intensity, perceived location, perceived quality, and perceived naturalness of the stimulus on each trial (**Figure 1**).

**Figure 1:**
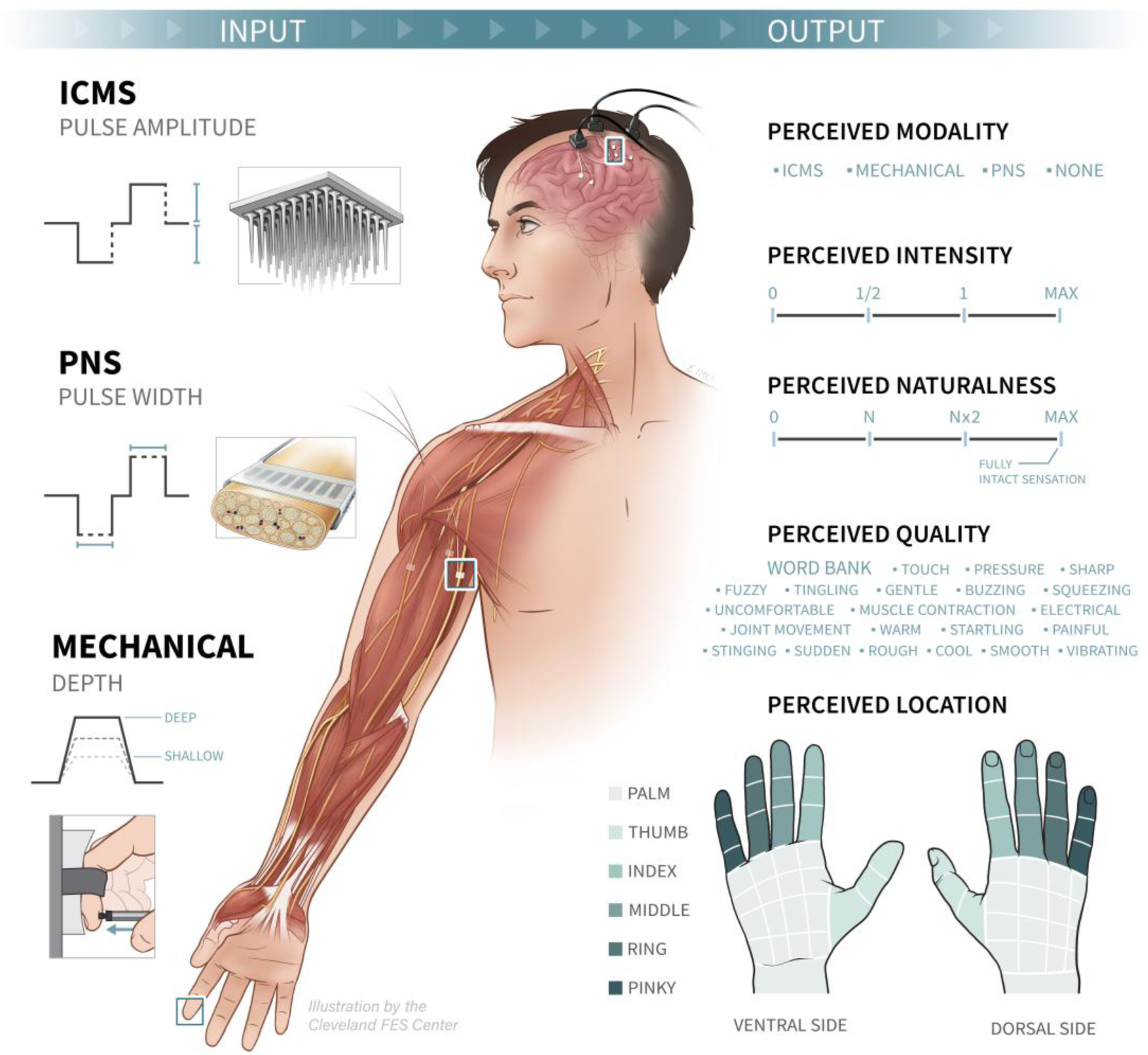
Participant with sensory-incomplete SCI compared PNS, ICMS, and mechanical touch. Sensory percepts evoked by intracortical microstimulation (ICMS), peripheral nerve stimulation (PNS), and mechanical indentation were compared in a human participant with sensory incomplete cervical-level SCI. ICMS was delivered via individual electrodes in a microelectrode array. PNS was delivered via individual contacts in a Composite Flat Interface Nerve Electrode (C-FINE). Mechanical indentation of the skin was delivered by a computer-controlled mechanical indenter. The stimulation level was set to 30%, 60%, and 90% of the sensory dynamic range for each stimulation modality. The stimulation parameter modified for ICMS was pulse amplitude (µA), for PNS was pulse width (µsec), and for the mechanical indenter was indentation depth (mm). The participant reported the perceived modality (ICMS, PNS, Mech, or none), intensity (numerical rating on an open-ended scale), naturalness (numerical rating on an open-ended scale), quality (set of descriptors from a word bank), and projected field (hand region) for each stimulation trial.

**Figure 2:**
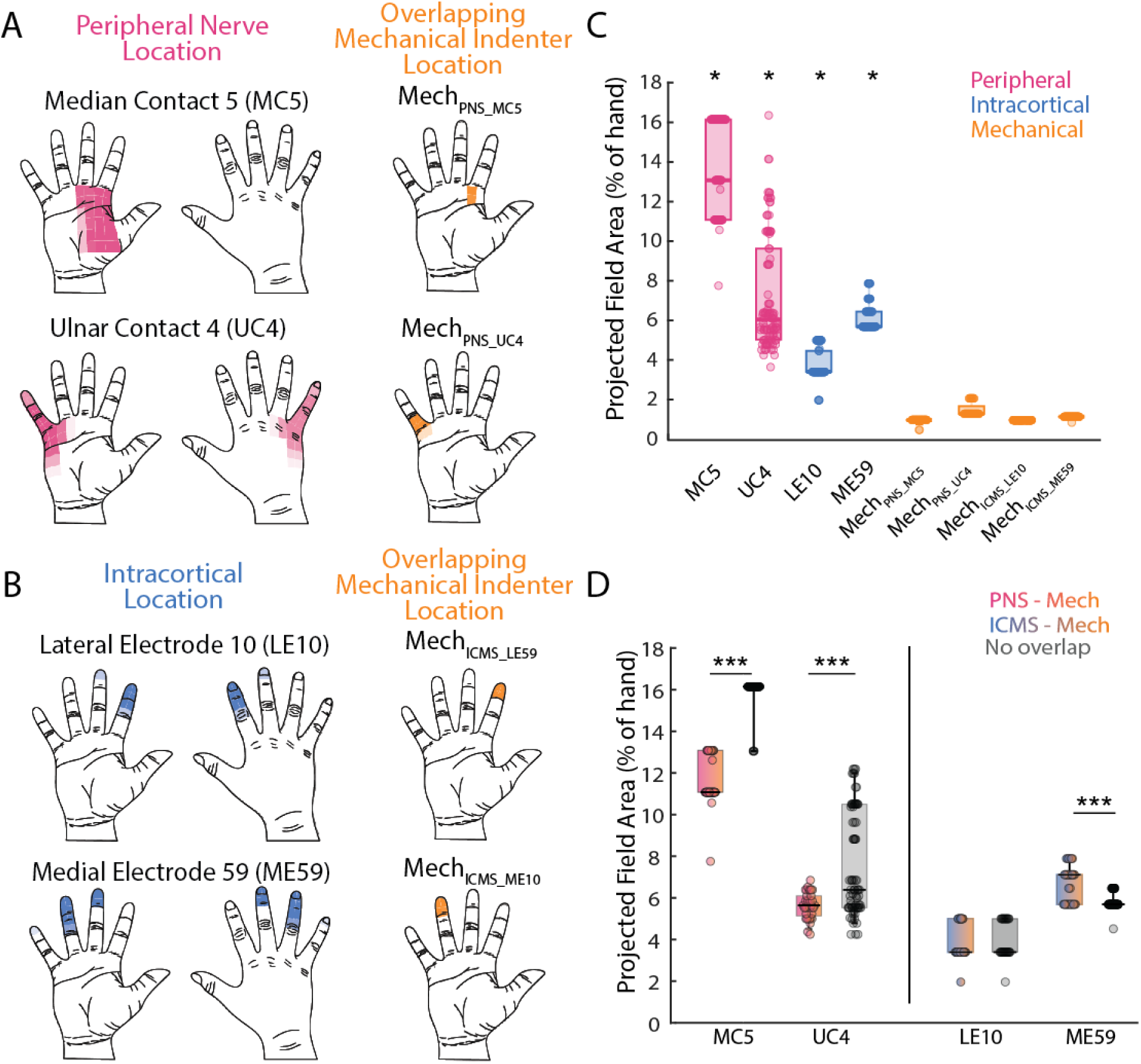
Sensory percept locations for ICMS, PNS, and mechanical stimulation. The projected fields reported for (A) PNS (median contact 5 and ulnar contact 4) and (B) ICMS (lateral electrode 10 and medial electrode 59) for all trials from one session across all stimulation levels. The opacity of the region indicates the frequency that region was reported across trials. Mechanical indentation (Mech) locations were chosen to spatially overlap with the percept locations for each PNS contact (Mech_PNS_MC5_ and Mech_PNS_UC4_) or ICMS electrode (Mech_ICMS_LE10_ and Mech_ICMS_ME59_) (A-B, orange regions). (C) The evoked percept location area (% of the total hand area) across all sessions for each contact/electrode/location tested. The * indicates significant differences (p<0.001) in percept size for that contact/electrode compared to all the other contacts/electrodes/ mechanical indenter positions. (D) Effect of overlap in mechanical indenter location with the corresponding PNS or ICMS evoked location on the projected field area compared to no overlap. The *** indicates significant differences at p<0.001. Note that all stimuli were delivered individually and that “overlap” refers to spatial overlap across trials within the session.

### PNS evokes larger percepts compared to ICMS and mechanical indentation

Both PNS and ICMS evoked sensation localized on the hand (**Figure 2**Error! Reference source not found.**A,B**). Overall, PNS produced significantly larger percept areas (percent of the whole hand area: 9.4 ± 4.1%) than ICMS (4.9 ± 1.4%), and both produced significantly larger percept areas than mechanical indentation (1.1 ± 0.22%) (One-way ANOVA: F(2,796) = 752.71, p<0.001, **Figure 2C**). The percept location areas evoked by each electrode of ICMS and each contact of PNS were significantly different compared to all other contacts/electrodes/locations across modalities (One-way ANOVA: F(7,791) = 660.55, p<0.001), but the mechanical indentation location areas were statistically indistinguishable from one another (**Figure 2C**). For PNS, MC5 evoked a significantly larger percept (13.81 ± 2.33%) than UC4 (7.28 ± 2.90%) (Tukey HSD post-hoc, p<0.001), and for ICMS, ME59 evoked a significantly larger percept (6.08 ± 0.67%) than LE10 (3.75 ± 0.76%) (Tukey HSD post-hoc, p<0.001). The percept sizes of the four mechanical indentation positions varied slightly but were not significantly different from one another (Mech_PNS_MC5_: 0.94 ± 0.12%, Mech_PNS_UC4_: 1.47 ± 0.35%, Mech_ICMS_LE10_: 0.96 ± 0%, Mech_ICMS_ME59_: 1.12 ± 0.03%). The perceived location and area were not affected by stimulation magnitude for PNS and mechanical indentation (**Supplementary Figure 2,3**). However, for ICMS lateral electrode 10, the perceived location became significantly larger by extending laterally to the middle finger for the high stimulation magnitude compared to the medium and low stimulation magnitude (One-way ANOVA: F(2,133) = 17.57, p<0.001, **Supplementary Figure 3**).

Our percept location analyses thus far pooled data across all contacts/electrodes tested to determine the effect of stimulation modality on projected field area, but did not consider the relative locations of the evoked percepts while making these comparisons. We next sought to investigate if the projected field area would differ when the percept locations evoked by each stimulation modality were spatially disparate vs when the percepts evoked by ICMS or PNS overlapped with those produced by mechanical indentation stimulation of the hand. Note that for all sessions, only one stimulation modality was applied in each trial, and the “overlap” of the percept locations refers to the similarity in percept locations across trials within the session. We observed that in sessions in which the percept location from mechanical indentation spatially overlapped with PNS (PNS-Mech), the PNS-evoked sensation was significantly smaller and more focal compared to sessions in which none of the percepts evoked by the three stimulation modalities overlapped (No overlap) (**Figure 2D**). This trend occurred for both median contact 5 and ulnar contact 4 (Wilcoxon rank-sum: p<0.001). However, when the percept evoked by ICMS overlapped with the mechanical indenter position (ICMS-Mech), the percept size was significantly larger than when there was no overlap (No Overlap) for medial electrode 59 (Wilcoxon rank-sum: p<0.001, **Figure 2D**). No difference was observed for lateral electrode 10, and thus the effect of percept overlap was not as consistent for ICMS as for PNS.

### PNS produces more intense and consistent percepts than ICMS

We then compared the perceived intensity scores reported by the participant across stimulation modalities. Each session consisted of 4-5 blocks of trials randomized across all three stimulation modalities, and the participant’s intensity ratings were normalized to the within-block mean. A few catch trials were also presented where no stimulation was delivered to the participant to confirm that the participant could reliably detect the stimulation across the three modalities. PNS was perceived to have significantly higher intensity (1.26 ± 0.31) compared to ICMS (1.03 ± 0.40), mechanical indentation (1.04 ± 0.38), and the catch trials (0.02 ± 0.12) (One-way ANOVA, F(3,1890) = 611.15, p < 0.001, **Figure 3A**). In contrast, the perceived intensity of ICMS was similar to mechanical indentation (Tukey HSD post-hoc, p = 0.99). The perceived intensity increased monotonically with stimulation magnitude for all sessions and stimulation modalities (One-way ANOVA, p < 0.001) (**Figure 3B)**. The spatial overlap of the PNS or the ICMS evoked percept with the mechanical indenter location within a session did not have any consistent effect on the perceived intensity (**Supplementary Figure 4A,B**).

**Figure 3:**
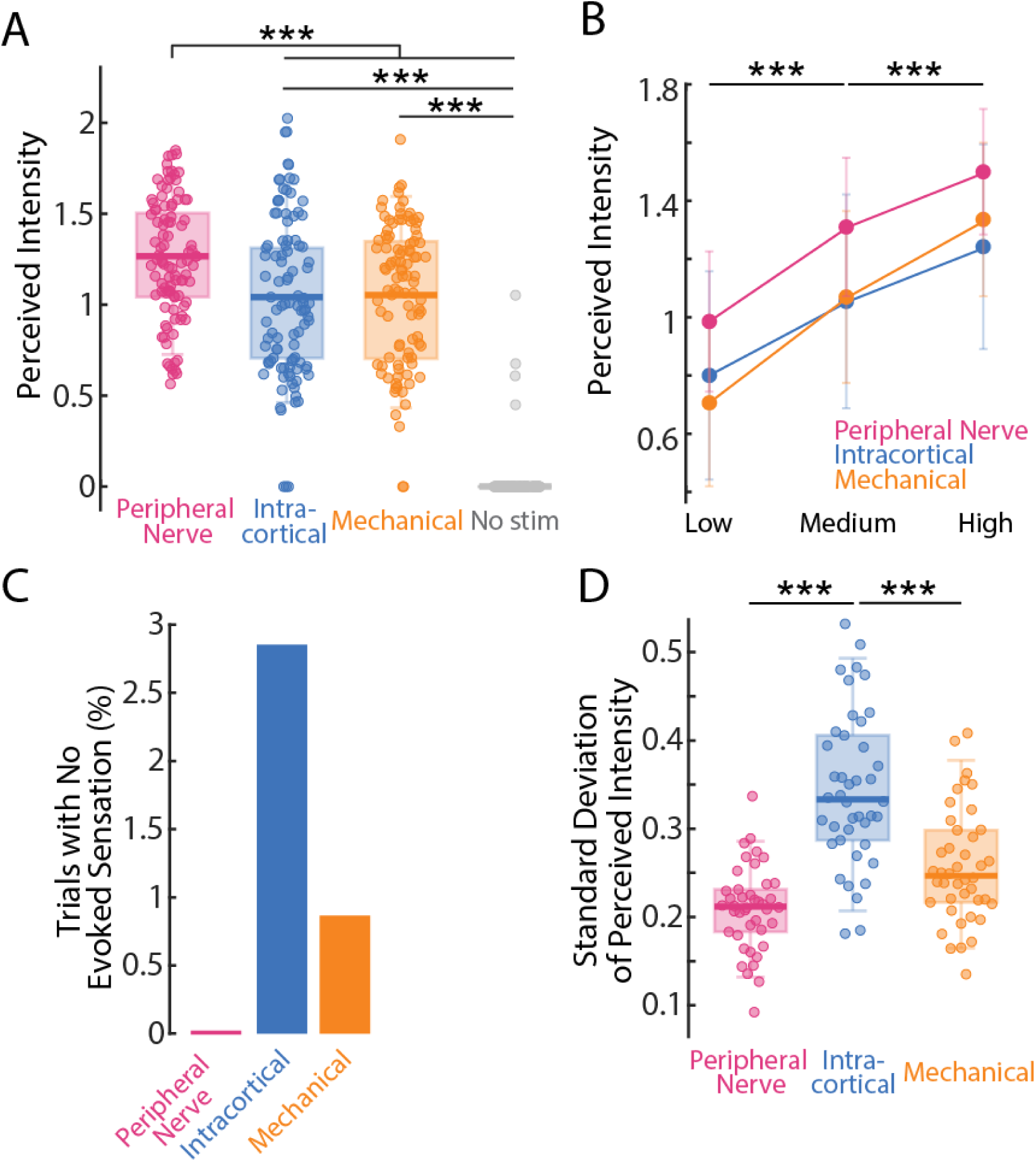
PNS evoked higher and more consistent perceived intensity than ICMS or mechanical indentation. A) The perceived intensities of the three stimulation modalities (PNS, ICMS and mechanical indentation (Mech)) and for the catch trials with no stimulation (shown in grey). For the stimulation modalities, data is combined from all three stimulation levels. B) The perceived intensity increased with stimulation magnitude (low, medium and high) for all three stimulation modalities. C) The percentage of trials for each modality out of all the trials where the participant did not detect a response. Sensation failed to be perceived in more trials for ICMS than PNS or Mech. D) The standard deviation of the perceived intensity within each stimulation level across all levels and sessions for the three modalities. All panels: *** indicates significance at p<0.001.

To ensure that the differences in perceived intensity across stimulation modalities were not driven by differences at a single stimulation level (e.g. effects restricted to the low stimulation level), we also compared the perceived intensities across modalities separately for each stimulation level (**Figure 3B**). Within each stimulation level, the perceived intensity of PNS was always highest (One-way ANOVA: low: F(2,569) = 43.4, p < 0.001; One-way ANOVA: medium: F(2,566) = 42.3, p < 0.001; One-way ANOVA: high: F(2,565) = 40, p < 0.001; Tukey HSD, p<0.001 for all levels). The perceived intensity of ICMS compared to mechanical indentation differed across stimulation levels: at the low stimulation magnitude, the perceived intensity of ICMS was significantly higher than mechanical indentation (Tukey HSD: p=0.006); at the medium stimulation magnitude, the perceived intensities were not statistically different (Tukey HSD: p = 0.875); and at the high stimulation magnitude, the perceived intensity of ICMS was significantly lower than mechanical indentation (Tukey HSD: p<0.005).

ICMS also failed to evoke sensation on the greatest percentage of trials (2.85%), compared to PNS (0%) and mechanical indentation (0.87%) (**Figure 3C, Supplementary Figure 5)**. The stimulation magnitudes where ICMS did not evoke any sensation were primarily low (2.32%) and medium (0.53%). Within sessions, the perceived intensities of PNS-evoked sensations were significantly more consistent across trials (i.e., lower standard deviation across trials of a given stimulation level within a session: 0.21 ± 0.05) compared to ICMS (0.34 ± 0.09) and mechanical indenter (0.26 ± 0.06) (One-way ANOVA: F(2,125) = 43.08, p<0.001, **Figure 3D**), and this trend held for all three stimulation magnitudes (**Supplementary Figure 6**). These results indicate that PNS evokes more reliable sensations and more consistent percept intensities than either ICMS or mechanical indentation.

### Perceived quality of ICMS-evoked sensation is similar to that of mechanical indentation

To assess the differences in the quality (or type) of evoked sensation across stimulation modalities, we asked the participant to describe the evoked sensation in each trial by selecting any number of descriptor words from a word bank (**Supplementary Table 2**). Since the mechanical indentation was applied to the hand, which is below the injury level, the participant was also asked to compare the quality of each evoked sensation to the quality of sensation felt during natural touch applied to the participant’s face (above the injury level). The natural touch sensation in the face was described as ”touch”, “pressure”, “gradual” and “gentle” (**Supplementary Figure 7A**).

The perceived quality of ICMS was more similar to mechanical indentation compared to PNS (**Figure 4A**). Both ICMS and mechanical indentation were reported in more than 99% of the trials as “touch”, “pressure”, and “tingling” sensation. In contrast, the PNS evoked sensation was reported as “sudden”, “buzzing”, “vibrating” and “electrical” in 100% of trials, and evoked sensations of “joint movement” in 87% of trials. The quality words selected for mechanical indentation on the participant’s finger mostly overlapped with natural touch on the face, except the participant did not feel “tingling” with above-injury natural touch but did for below-injury touch. ICMS-evoked sensation was also reported as “sudden” and “squeezing” in 100% of trials, but these descriptors were typically not reported for mechanical indentation on the hand or natural touch on the face.

**Figure 4:**
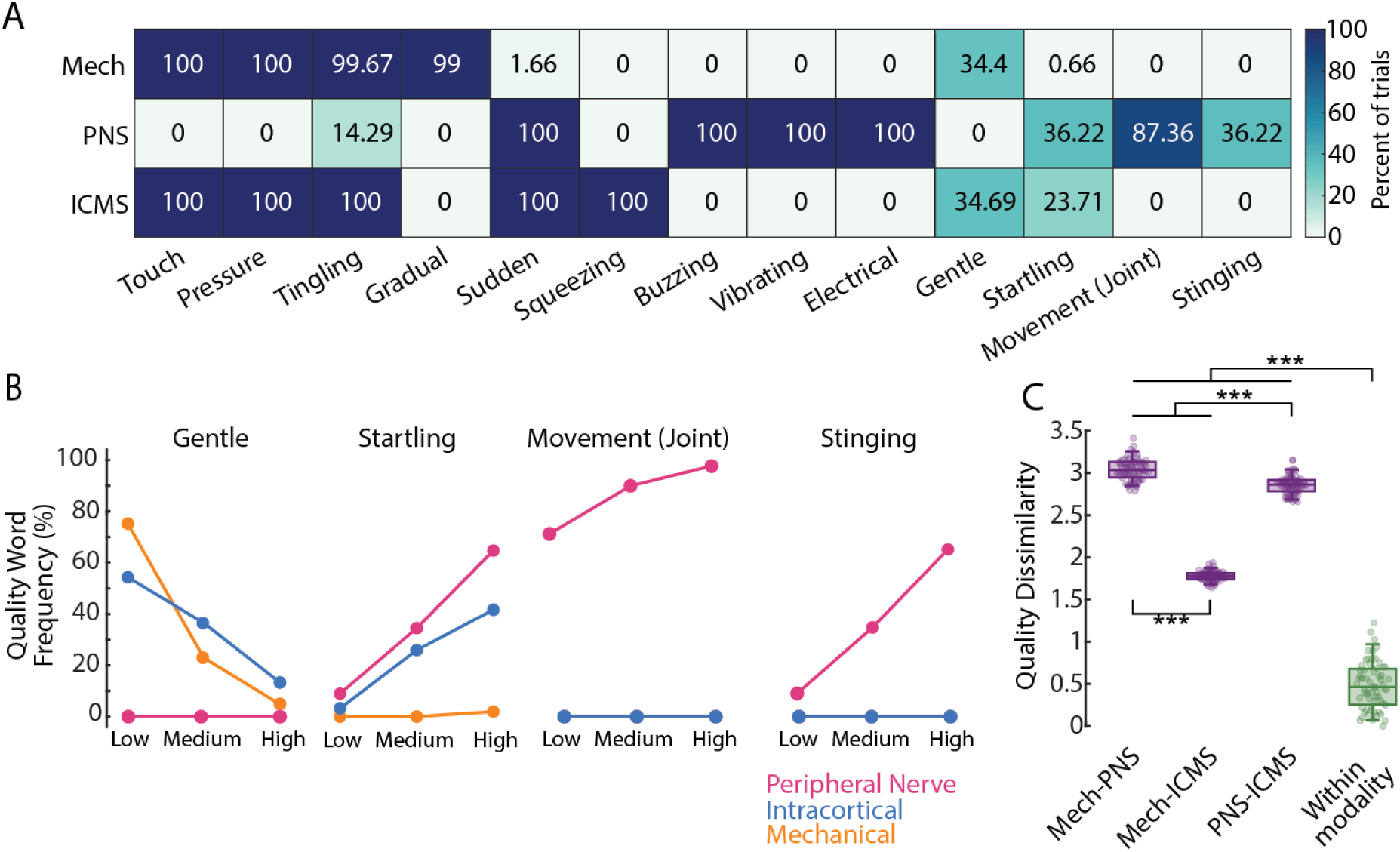
**The perceived quality of ICMS was more similar to mechanical indentation compared to PNS.** A) The frequency of quality descriptor words selected for each stimulation modality. Data is shown as the percentage of trials across all sessions and stimulation levels in which each word was reported for each modality. B) Stimulation magnitude (low, medium, high) impacted the frequency that some quality words were selected by the participant to describe the perceived sensation. Only those quality words that exhibited large changes across stimulation levels for any stimulation modality are plotted. C) Quality dissimilarity between mechanical indentation and PNS trials (Mech-PNS), between mechanical indentation and ICMS trials (Mech-ICMS), between PNS and ICMS trials (PNS-ICMS), and across stimulation levels within a stimulation modality (Within Modality). The *** indicates significant differences between the groups (p<0.001).

With increases in stimulation magnitude, ICMS felt less ‘gentle’ and more ’startling’ **(Figure 4B)**. For PNS, increases in stimulation magnitude increased ‘startling’ and ‘stinging’ sensations, as well as making the sensations feel more like ‘joint movement’ **(Figure 4B)**. Mechanical indentation and natural touch felt less ‘gentle’ with increasing stimulation magnitude (**Supplementary Figure 7**).

To quantitatively compare the degree of similarity in perceived quality among the stimulation conditions, we then transformed the descriptor frequencies into dissimilarity ratings based on Euclidean distance, as described in our prior work^12^. The dissimilarity in perceived quality between ICMS and mechanical indentation (0.56 ± 0.01) was significantly lower than between PNS and mechanical indentation (0.95 ± 0.03) and between PNS and ICMS (0.89 ± 0.03) (One-way ANOVA: F(2,24) = 578.76, p<0.001) (**Figure 4C**).

This indicates that the perceived quality of ICMS was closer to that of mechanical indentation than PNS was to mechanical indentation. The spatial overlap of the PNS- or ICMS-evoked percept with the mechanical indenter position within a session did not affect the reported quality for any of the three modalities (**Supplementary Figure 4E,F**).

### Perceived Naturalness of ICMS is Higher than PNS

The participant also reported the perceived naturalness for each stimulus. The participant was instructed to compare the sensation evoked by the stimulus to natural touch applied to their face, which they ascribed the highest rating (10). The perceived naturalness of PNS (1.19 ± 0.63) was by far the lowest, compared to ICMS (6.68 ± 0.64) and mechanical indentation (7.60 ± 0.72). These differences across stimulation modalities were statistically significant (One-way ANOVA: F(2,1605) = 15414.13, p < 0.001), and each stimulation modality had significantly lower naturalness ratings than natural touch applied to the face (one-sample Wilcoxon signed-rank test, p<0.001) (**Figure 5A**). Although the perceived naturalness of ICMS was closer to that of mechanical indentation on the hand, it was still significantly lower (posthoc Tukey HSD, p < 0.001). The spatial overlap of the PNS- or ICMS-evoked percept with the mechanical indenter location within a session did not have any consistent effects on the perceived naturalness for any of the three modalities (**Supplementary Figure 4C,D**). Although there were significant differences in naturalness across modalities, within each modality, we saw consistent monotonic decreases in naturalness with increasing stimulation magnitude (One-way ANOVA: p < 0.0001, Tukey HSD: p < 0.001 (**Figure 5B**).

**Figure 5:**
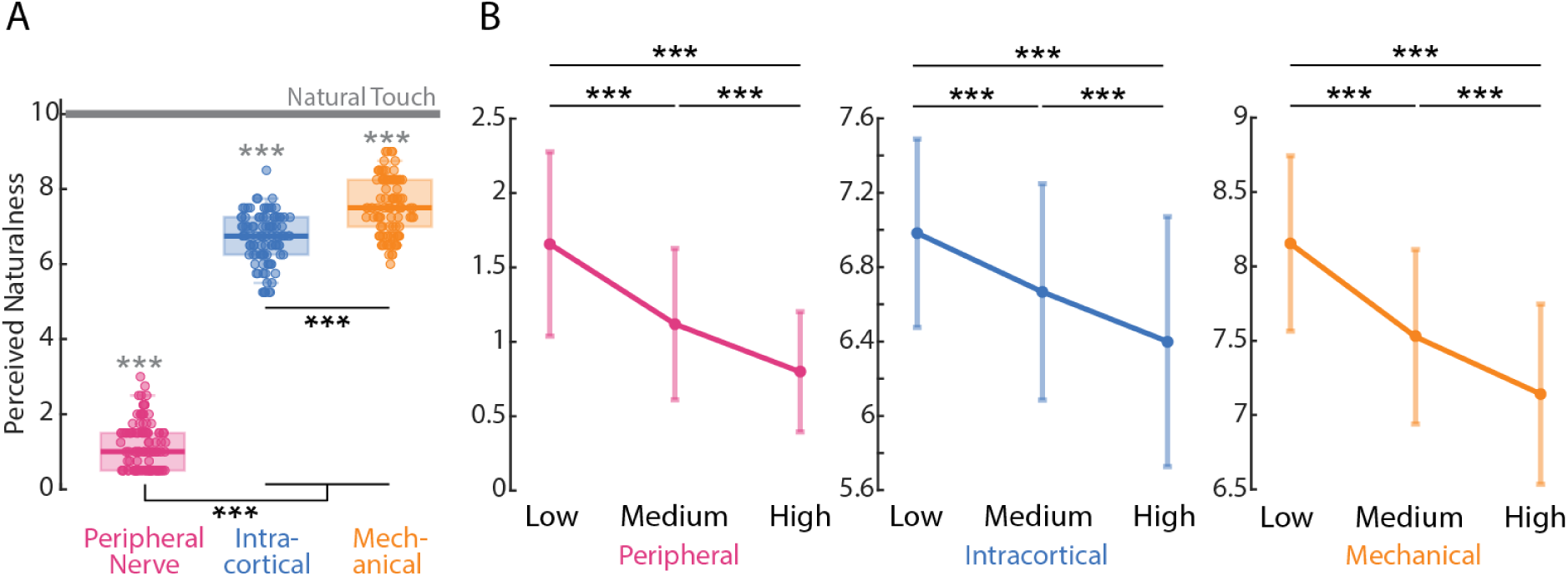
Perceived Naturalness is Higher for ICMS than PNS. A) The perceived naturalness reported for the three stimulation modalities (PNS, ICMS, Mechanical), with data combined across all stimulation levels and sessions. The *** (marked in black) indicates significant differences for the indicated stimulation modality compared to the other modalities (Tukey HSD, p<0.001). The grey line indicates the perceived naturalness reported for natural touch applied above injury to the face. The *** (marked in grey) indicates significant difference between the indicated stimulation modality and natural touch (one-sample Wilcoxon, p<0.001). B) Perceived naturalness decreased with increasing stimulation magnitude (low, medium, and high) for all three stimulation modalities. *** indicates p<0.001, Tukey HSD. The y-axis limits were adjusted for each modality to highlight the differences across stimulation magnitudes.

### Perceived Naturalness is Better Predicted by Perceptual Metrics than Stimulation Conditions

The perceived naturalness of an evoked sensation may be an important aspect of its acceptability in a neuroprosthetic application, but the definition of “naturalness” remains controversial in the field of sensory neuroprostheses. We sought to shed light on this issue by relating the perceived naturalness of stimulation-elicited sensations to other aspects of the sensory experience. Given that we observed robust effects of our stimulation variables (stimulation modality, stimulation magnitude) on perceived naturalness (**Figure 5**), as well as effects of the stimulation variables on the other measured perceptual variables (perceived intensity, quality) (**Figure 3-4**), we sought to determine the extent to which these perceptual variables co-varied across trials.

We found that for all stimulation modalities, the perceived naturalness was negatively correlated to perceived intensity (linear regression: R^2^ = 0.67, p<0.001 for PNS, R^2^ = 0.56, p<0.001 for ICMS, R^2^ = 0.53, p<0.001 for mechanical indenter). This negative relationship had approximately the same slope for all stimulation modalities (linear regression coefficients: −0.36 for PNS, −0.28 for ICMS and −0.34 for mechanical indentation) (**Figure 6A**). We also performed a similar analysis to determine the correlation between perceived naturalness and perceived quality. Because quality occupied a multidimensional space, we performed a multiple linear regression to predict the average perceived naturalness of each condition for each session from the quality descriptor space for that condition/session. We found that the perceived quality very strongly predicted the perceived naturalness (R^2^ = 0.99 ± 0.004, p<0.01) (**Figure 6B**). Therefore, both perceptual variables (perceived intensity and quality) and stimulation magnitude (**Figure 5**) determine the perceived naturalness, but it’s unclear whether the stimulation magnitude has a larger impact over perceptual variables on perceived naturalness. To determine the extent to which this relationship between perceptual variables was influenced by the stimulation magnitude (i.e., the stimulation variable rather than the perceptual variable), we first regressed the perceived naturalness with stimulation magnitude, then regressed the residuals with the perceived intensity. This analysis yielded a significant relationship between the perceived naturalness and perceived intensity even after accounting for the stimulation magnitude (Linear regression: R^2^ = 0.28 for PNS, 0.41 for ICMS, 0.2 for mechanical indentation, p<0.001 for all).

**Figure 6:**
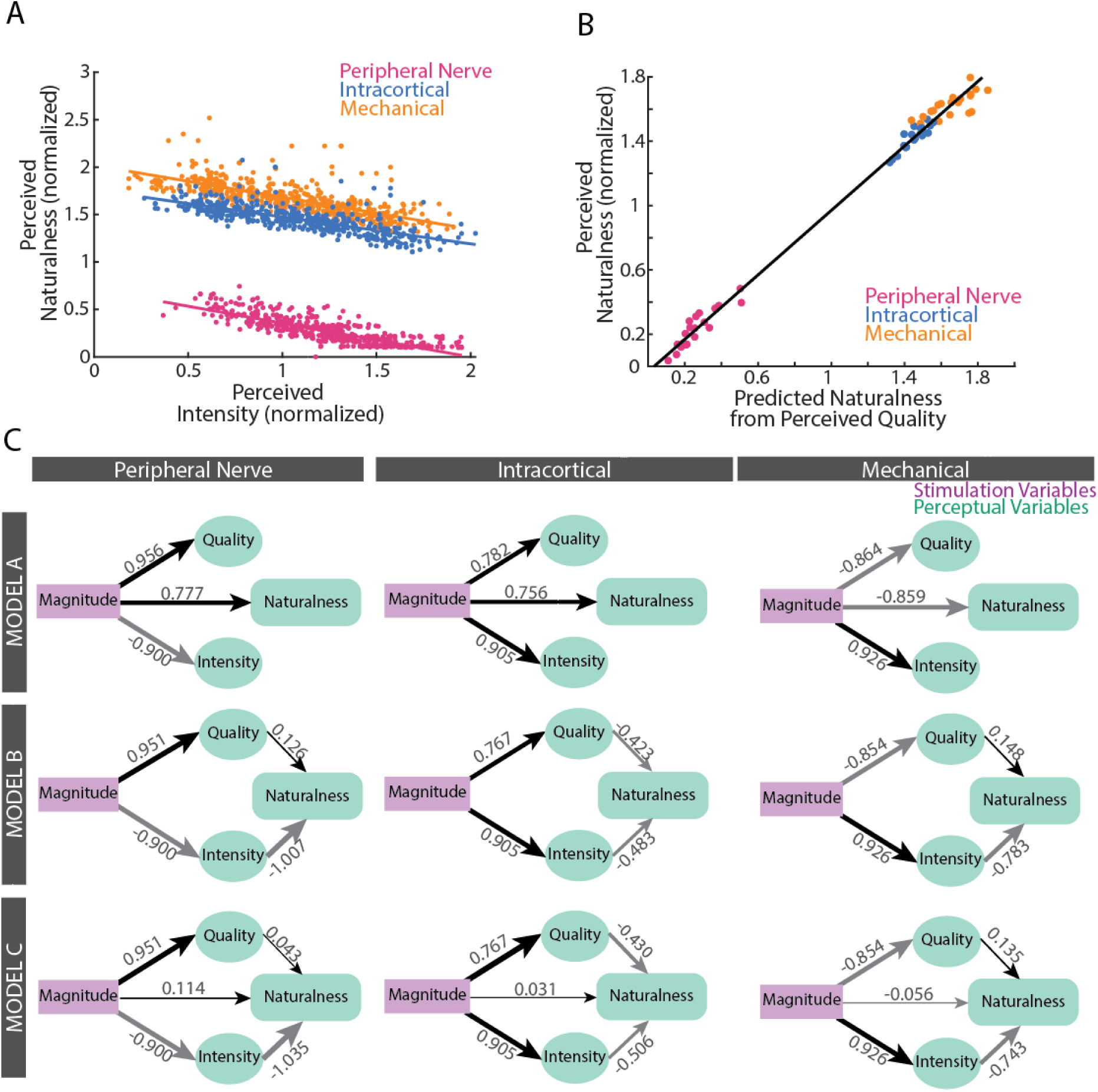
Perceived Naturalness is Better Predicted by Perceptual Variables than by Stimulation Variables. A) Normalized perceived naturalness was highly correlated to normalized perceived intensity for all three stimulation modalities. (B) Relationship between the perceived naturalness reported by the participant (y axis) and the predicted naturalness from a linear regression of perceived quality (x axis) across all three stimulation modalities. (C) PLS-SEM depicting three potential relationships among the stimulation (purple) and perceptual (teal) variables in this study for the three stimulation modalities. In Model A, the stimulation magnitude predicts the perceptual variables (quality, intensity, and naturalness), and there is no interaction among perceptual variables. In Model B, the stimulation magnitude directly predicts perceived quality and intensity, which serve as predictors of perceived naturalness. In this model, we assume that the stimulation variables only indirectly influence perceived naturalness through the mediating effects of the other perceptual variables. In Model C, perceived naturalness is predicted by both the stimulation magnitude and the other perceptual variables. The normalized coefficient for each relationship is indicated by the overlaid values, as well as the arrow width. Black arrows denote positive coefficients, and grey arrows denote negative coefficients.

Given that these analyses demonstrate that perceived naturalness can be predicted on the basis of either stimulation variables or perceptual variables, the question is which set of variables best predicts perceived naturalness: stimulation variables or perceptual variables? If perceived naturalness is best predicted by perceptual variables, this would suggest that naturalness is a higher-order perceptual dimension that the participant constructs by combining judgments in other primary perceptual dimensions (such as intensity, quality, location, and/or timing). To answer this question, we constructed a series of partial least squares structural equation models (PLS-SEM)^66,67^ for each stimulation modality where naturalness was predicted by stimulation magnitude and/or other perceptual variables (i.e. quality and intensity) (**Figure 6C**). We compared two models. Model A assumed that naturalness, as well as perceived intensity and quality, depended entirely on stimulation magnitude. This model represents the idea that perceived naturalness is an independent perceptual dimension that does not depend on perceived intensity or quality. Model B assumed that perceived intensity and quality mediate the relationship between stimulation magnitude and perceived naturalness. This model represents the idea that perceived naturalness is not independent of other perceptual variables and instead is a higher order dimension that is innately linked to judgments in the other perceptual dimensions. Model C is a “control” model for comparison purposes. In Model C, Models A and B were combined such that perceived naturalness was predicted by both the stimulation and the perceptual variables. Since Model C included weightings for all inter-variable relationships, we expected that it would yield the best fit to the data and thus represents the “gold standard” for model fit to which to compare the fits of Models A and B. Model C was also used to compare the regression coefficients to determine the relative strengths of the relationship between stimulation variables and naturalness compared to the relationship between perceptual variables and naturalness.

Overall, Model B outperformed Model A in both R-squared and standardized root mean square residuals for all three stimulation modalities (**Table 1**). This indicates that naturalness was better modeled as a higher dimension that depended on other perceptual variables than as an independent concept that was judged separately from the other perceptual variables (**Table 1**). The standardized root mean square residuals for all the models across the three stimulation modalities were less than 0.08, thereby indicating a good model fit^49^. Furthermore, the standardized root mean square residuals for Model B & C were less than Model A. The R^2^ value of naturalness was higher for Model B & C compared to Model A for all the three stimulation modalities.

**Table 1:** Metrics of model performance for the three PLS-SEM of perceived naturalness.

| Standardized root mean square residuals |  |  |  |
| --- | --- | --- | --- |
|  | Peripheral Nerve | Intracortical | Mechanical |
| **Model A** | 0.046 | 0.066 | 0.05 |
| **Model B** | 0.036 | 0.051 | 0.04 |
| **Model C** | 0.036 | 0.051 | 0.04 |
| $R^2$ (Naturalness) | | | |
|  | Peripheral Nerve | Intracortical | Mechanical |
| **Model A** | 0.60 | 0.57 | 0.74 |
| **Model B** | 0.80 | 0.72 | 0.83 |
| **Model C** | 0.80 | 0.72 | 0.83 |

In addition, the absolute value of the normalized regression coefficients in Model C give us insight into the relative contributions of each variable in predicting perceived naturalness. For all stimulation modalities, the perceived variables had higher regression coefficients in Model C than those for stimulation magnitude (**Figure 6C**). Specifically, the coefficient for the relationship between perceived intensity and naturalness was more than 9 times the coefficient for the relationship between stimulation magnitude and naturalness, and this trend held for all three stimulation modalities. This indicates that compared to stimulation magnitude, the perceptual variables much more strongly predicted perceived naturalness. Interestingly, the regression coefficients for the relationship between perceived intensity and naturalness and between perceived quality and naturalness were approximately the same for the ICMS model, but for the PNS and Mechanical models, the coefficients for the relationship between perceived intensity and naturalness were ∼5-24x larger than those for the relationship between perceived quality and naturalness. We also observed that for all three stimulation modalities, the perceived intensity and quality were strongly predicted by stimulation magnitude in Model C, as evidenced by the relatively large regression coefficients for these relationships.

Taken together, the results of the regression analyses, the better fit of Model B compared to Model A, and the regression coefficients in Model C all support the conclusion that the perceptual variables (quality and intensity) more strongly predicted perceived naturalness than did the stimulation variables. These findings suggest that perceived naturalness is a higher order perceptual dimension that depends on the participant’s experiences and judgments of other perceptual dimensions.

## Discussion

Many research groups and companies around the world are seeking to develop bidirectional brain computer interfaces to restore sensory and motor function to people with injuries leading to paralysis or limited upper extremity function. Towards delivering effective sensory feedback for these neuroprosthetic systems, for the first time, we directly compared evoked sensations from cortical and peripheral stimulation in an individual with incomplete SCI. While both stimulation modalities have been shown to evoke sensations on the hand in prior studies, the perceptual differences between PNS and ICMS have been difficult to ascertain because of differences in the patient populations studied for each modality, the heterogeneity in methods used to measure the percepts, and variance across studies, subjects, and stimulation sites^3,11,12,15,50^. Direct comparisons of the evoked percepts from PNS and ICMS had never been performed because no individual had devices capable of delivering both stimulation modalities. However, our study participant had both intracortical arrays and peripheral nerve electrodes implanted through his participation in the ReHAB clinical trial (ClinicalTrials.gov #NCT03898804), and this enabled us to directly compare the sensations produced by PNS and ICMS^36^.

This is also the first study in which PNS was explored as an approach for sensory restoration in individuals with sensory-incomplete SCI. Prior studies of sensory restoration after SCI have focused on brain stimulation to avoid the potential effects of disruption of the sensory information travelling to the cortex caused by the SCI. However, many people with SCI have at least partially preserved below-injury sensory function^51^, and the extent to which damage to the spinal pathway affects the perception of below-injury PNS has never been studied before. In this study, we showed as a proof-of-concept that an individual with AIS-B SCI can reliably detect, perceive, and describe sensations evoked by both peripheral nerve and cortical stimulation. This study directly and quantitatively compared the perceptual dimensions of PNS- and ICMS-evoked sensation and opens the door for further development of PNS as a viable sensory feedback modality for people with SCI.

We observed that compared to PNS-evoked percepts, the ICMS-evoked percepts were closer to those evoked by mechanical indentation applied to the hand in both perceived quality and naturalness. A possible explanation for why ICMS feels qualitatively more similar to mechanical touch could be related to differences in somatosensory information representation in the cortex versus the periphery. In the periphery, different afferent types, such as slowly adapting (SA) or rapidly adapting (RA) fibers, precisely convey information about different aspects of natural touch stimuli via specific temporal firing patterns^52^. Past studies have shown that multiple peripheral inputs converge at multiple stages along the sensory neuraxis before S1 and are processed to extract relevant perceptual features^53–55^. Therefore, ICMS in area 1 may evoke more natural sensation because it recruits neurons that represent higher-order perceptual features that have already been pre-processed at other cortical and subcortical structures. Additionally, modeling studies have shown that PNS synchronously recruits mixed populations of SA and RA fibers in the periphery,^56,57^ and these evoked firing patterns differ from those produced in peripheral afferents by natural touch^58–60^. Because peripheral neural signals convey basic properties of the touch stimulus, it is possible that any mismatch in firing pattern in the periphery due to PNS has a larger impact on the perceptual experience than any mismatch in firing pattern in the cortex due to ICMS. One important caveat is that our study was conducted with simple stimulation paradigms for both PNS and ICMS that made no attempt to reproduce or mimic the natural firing patterns of touch. Many studies are investigating biomimetic stimulation paradigms as an approach to improve the naturalness and/or quality of sensations evoked by neurostimulation^17,39,41,61^. Therefore, future work will replicate our study with biomimetic stimulation to determine how the stimulation paradigm impacts the differences in quality and naturalness between ICMS and PNS observed here.

One possible explanation for the difference in the perceptual properties of ICMS and PNS is that the SCI disrupted the afferent signals from the periphery during PNS. If the SCI altered the PNS-evoked sensation, we would expect that it would have similarly impacted the mechanical indentation-evoked sensation, which was also applied to the hand below the injury level. Indeed, we observed that mechanical indentation applied to the hand (below injury) evoked more tingling-type sensations that were lower in naturalness than touch applied to the face (above injury), demonstrating that the SCI did impact the participant’s perception of the below-injury peripheral inputs. Such differences in the naturalness of tactile sensation in SCI-impaired versus non-impaired skin regions align with previous studies^61^. However, if the SCI was the primary determinant of the quality and naturalness of PNS-evoked sensation, we would have expected that both mechanical indentation and PNS would have had similarly low naturalness levels and similar unnatural qualities (i.e., that mechanical indentation-evoked sensation would have been more similar to PNS than to ICMS). This is not what we observed. Instead, the quality and naturalness of ICMS was most similar to mechanical indentation, and PNS was much less natural and distinctly more dissimilar in quality than both ICMS and mechanical indentation. Therefore, while the impact of the SCI cannot be excluded, other factors, such as the firing patterns evoked in the stimulation-recruited neural populations, also likely played a role in the perceptual differences between ICMS and PNS.

The sizes of the ICMS-evoked percepts were also smaller and more focal than the percepts evoked by PNS. This finding is consistent with prior studies, which found that ICMS projected fields can be highly focal (fingertip size or smaller)^20,61,62^ or encompass several finger regions^15,36,61,62^. PNS projected fields in prior studies ranged from the size of a fingertip or smaller^11,13,63^ to entire fingers^11,40^ or large swathes of the palm^13,28,40^, depending on the type of neural interface and the stimulation parameters^11^. The size of the projected field from any stimulation modality depends on the somatotopic organization of the neural structure and the number of neurons recruited from the stimulation^15,37^. ICMS is believed to directly recruit cortical neurons up to 2 mm away from the electrode, with the strongest activation <500 µm^64–66^, and can also sparsely recruit neurons by activating passing axons^66,67^. The region of neurons directly recruited by ICMS is smaller than the distance between the somatotopic organization of the fingers in Brodmann’s area 1, which ranges from 3 to 7 mm apart^68^. Therefore, ICMS likely recruits neurons projecting to regions smaller than a finger and hence can evoke highly focal percepts. Although PNS can recruit sub-fascicular regions of the nerve^35^, depending on the stimulation parameters, the somatotopic organization in the fascicles at the electrode location likely impacts the size of the projected field from PNS. Prior anatomical and histological studies revealed that the neurons from a given skin area intermingle across fascicles as they travel proximally ^69^. Thus, recruiting even part of a single fascicle in a proximal position could lead to activation of afferents from a wide area of the hand and arm. In this study, the C-FINEs were implanted in the most proximal location possible - just after the branch point from the brachial plexus. Hence, it is likely that the afferents were more disorganized at this proximal level, resulting in larger, more diffuse percepts from PNS than could be achieved if the peripheral electrodes had been implanted more distally.

We found that PNS-evoked percepts were more reliably detected and more consistent in reported intensity than the ICMS-evoked percepts. The participant was also least likely to confuse PNS with any of the other stimulation modalities, including catch trials (i.e., no stimulation). Following the same rationale that we posited for the differences in naturalness between ICMS and PNS percepts, the higher reliability of PNS compared to ICMS could also relate to the nature of the information represented by peripheral vs cortical neurons. Peripheral afferents precisely convey information about touch stimuli from the external world to the brain^52^, and due to the high likelihood of variability in external conditions, downstream perceptual processes may be relatively robust to small variations in peripheral neural firing that the system has learned do not convey meaningful information about the external stimuli. Thus, if PNS recruited peripheral afferents in approximately the same way in each trial, it is reasonable that the variability in the perceptual experience would be low. In contrast, assuming ICMS activates higher-order neurons that represent perceptual features^53,54^, small variations in ICMS-recruited neural activity from trial to trial may indicate a large change in its represented feature(s). In addition, these cortical neurons may also be influenced by brain state and/or inputs from many other cortical areas^70^, leading to large variations in excitability or responsiveness to ICMS across trials. Thus, these factors may have contributed to the larger variability in perceptual experience across ICMS stimuli.

The perceived intensity for PNS was also higher with lower variability than both ICMS and mechanical indentation. Despite our systematic procedure to find an equivalent sensory dynamic range for each stimulation modality, the perceived intensity of PNS was reported to be greater on average than ICMS or mechanical indentation. Because we selected stimulation magnitudes at specific percentages of the sensory dynamic range (30%, 60%, 90%), this difference in average perceived intensity could have arisen if the function relating PNS stimulation magnitudes to intensity level was more logarithmic than that of ICMS or mechanical indentation. Future work could test this hypothesis by conducting a magnitude estimation task with a set of stimulation magnitudes constructing more complete models relating stimulation level to perceived intensity for each stimulation modality. The higher average perceived intensity of PNS may also have been impacted by differences in adaptation rates across the stimulation modalities^50,71^, and/or that the participant may have had higher tolerance to stimulation, and thus a higher maximum comfortable limit, for PNS than for the other modalities. Given that the perceived intensities across all stimulation levels were shifted upward for PNS, the effective sensory dynamic range for PNS was likely wider than for ICMS or mechanical indentation.

Taking together, our findings demonstrate that there are benefits to both ICMS and PNS, and thus both stimulation approaches have merit for bidirectional sensorimotor neuroprostheses. However, given their specific strengths and weaknesses, each stimulation approach may be best suited for different purposes. Because of ICMS’s enhanced naturalness and quality, ICMS may be the best approach to restore the affective aspects of touch^72^, in which the naturalistic quality of the sensation could be imperative for emotional and social connection to other people. ICMS may also be better able to communicate complex object properties, such as compliance or texture, to a neuroprosthesis user. PNS, in contrast, may be the best stimulation modality to convey consistent, reliable sensation during functional task performance. Past studies have shown that a stimulus with higher perceived intensity has a longer adaptation time^71^, allows for higher resolution feedback, and improves functional task performance^28^. During a functional task, the mental effort required to control a motor neuroprosthesis, the cognitive demands of the task itself, as well as the barrage of other incoming visual and auditory signals, could alter or impair the somatosensory information^73,74^ provided by sensory neurostimulation^75^. The stronger and more salient information provided by PNS could be critical to successfully interpreting and utilizing the somatosensory feedback to effectively complete the task. Future work should compare functional task performance with feedback provided by each of the stimulation modalities to determine whether sensory feedback provided via PNS is in fact more effective than that provided via ICMS.

One of the primary focus areas of the sensory neuroprosthesis field in recent years has been improving the naturalness of artificially-evoked sensation^7,76–79^. Prior studies have reported how perceived naturalness changes with stimulation variables like stimulation magnitude^80^ and spatiotemporal patterning^17,39,41^. We were curious to see if perceived naturalness was also dependent on other perceptual dimensions like intensity and quality. Interestingly, the PLS-SEM models support the idea that naturalness is not an independent perceptual dimension from intensity and quality, but rather a higher cognitive construct that depends more on these other perceptual dimensions than on stimulation variables. This interpretation is further supported by the similarities in the relationships observed between the perceived intensity and naturalness of the sensations across all stimulation modalities. This indicates that while varying stimulation parameters has been shown to improve the naturalness of an evoked sensation^17,41^, the other perceptual dimensions of a percept, such as its intensity and quality, play a major role in shaping the participant’s interpretation of the perceived naturalness of the sensation. This finding suggests that perceived naturalness may be governed by the participant’s views or beliefs about what kinds of sensory experiences should be deemed ‘natural’. Thus, a particular stimulation modality or paradigm that is reported as ‘natural’ by one individual might not be judged as ‘natural’ across participants. This indicates that neurostimulation paradigms to enhance perceived naturalness may need to be tailored for each individual based on their preferences and beliefs.

This study presents a unique comparison of cortical and peripheral stimulation approaches to restore somatosensation in people with sensory-incomplete SCI. Our results show that ICMS feels more similar to mechanical indentation in quality and naturalness, indicating ICMS may be a better stimulation modality to restore affective touch or complex information about object properties. On the other hand, PNS evoked stronger and more reliable percepts, indicating PNS may be a better modality for delivering real-time sensory feedback during closed-loop functional tasks. Our analyses also revealed that perceived naturalness is a higher perceptual dimension that depends more heavily on perceptual variables than stimulation variables, which supports the importance of patient-specific stimulation paradigms for improving sensation naturalness. Although our findings are limited to one participant, this study highlights for the first time that PNS is a viable option for sensory restoration for individuals with sensory-incomplete SCI, and thus PNS could be used instead of, or in conjunction with, ICMS as a feedback approach for brain computer interfacing neuroprostheses. In the future, delivering ICMS and PNS simultaneously will be explored to see if the brain integrates reliable information from PNS with the natural quality of ICMS, resulting in expanded capabilities and improved overall experience of sensation restored via neurostimulation.

## Methods

### Research Participant

One participant with an AIS-B C3-C4 spinal cord injury was enrolled in the Reconnecting the Hand and Arm to the Brain (ReHAB) clinical trial^36^ (ClinicalTrials.gov #NCT03898804). The participant’s intact residual sensation was evaluated using the Semmes-Weinstein monofilament^81^ and the 2-point discrimination^82^ clinical tests (Supplementary Table 1). As part of the brain computer interfacing system received in the ReHAB study, the participant was implanted with two 64-channel (8x8) microelectrode arrays (Blackrock Neurotech, Salt Lake City, Utah, USA) in the hand area of the left primary somatosensory cortex (Brodmann’s area 1) to provide ICMS. As part of the functional electrical stimulation system received in the ReHAB study, the participant received 16-channel composite flat interface nerve electrodes (C-FINEs) that were placed around the right median and ulnar nerves. These C-FINEs were used to provide sensory PNS. More details about the implanted devices and surgical approach for this participant can be found in Herring et al., 2024^36^.

### Stimulation Parameters

ICMS was delivered by a Cerestim C96 multichannel microstimulation system (Blackrock Neurotech, Salt Lake City, Utah, USA) through an analog 128-channel stimulation cable to individual electrodes in the two microelectrodes implanted in Brodmann’s area 1. PNS was delivered by a Universal External Control Unit (UECU) through individual contacts in the C-FINEs on the median and ulnar nerves (Technical Development Laboratory, Cleveland, OH, USA)^83^. Both ICMS and PNS stimuli were cathodal phase first, current-controlled, charge-balanced pulse trains^11,15^ delivered at 50 Hz for 1 second duration. For ICMS, the cathodal phase pulse width was 200 µs and the pulse amplitude of the cathodal phase was varied over a range from 0 to 100 µA to modulate the sensation intensity^15^.^15^ For PNS, the pulse amplitude was set on a per-day basis to the minimum level found to reliably evoke sensation (typically 0.5 mA or 0.6 mA)^11^. PNS pulse width varied over a range of 50 to250 µsec to modulate the sensation intensity^11^. The values of pulse amplitude and width of each stimulus were balanced such that the charge injected per phase was under the Shannon safety limits^11,15,84–86^.

Mechanical indentation stimulation was applied using a computer-controlled tactile linear actuator (SMAC LCA25-025-31, Triad Technologies, Akron, OH). The mechanical indentations were 1-second-long symmetric trapezoidal indentations presented to the ventral side of RP1’s hand with a 3 mm diameter blunt probe^87–89^. The indentation depth was varied over a range of 0 to 5 mm to modulate the perceived intensity^90^. The onset and offset rate were 40 mm/sec^87^. The probe tip was pre-indented at the beginning of each block to account for small fluctuations in the hand position relative to the mechanical indenter^87^. The stimulation shape, timing, and parameters were based on prior human and NHP literature^87–91^, although the indentation depths were increased slightly such that the participant could reliably detect tactile indentation despite his SCI.

### Experimental Design

The sensory dynamic range was defined as the range of stimulation magnitudes between the participant’s threshold and maximum comfortable perceived intensity. The threshold was determined using a staircase with reversals method where the stimulation magnitude was increased by a fixed step size until the participant detected a sensation, then decreased by a smaller step size until the participant stopped reporting a detected sensation and then reversed three more times with the same logic^92^. The threshold was computed by averaging the stimulation magnitudes at each reversal together^92^. The maximum comfortable perceived intensity was determined by incrementing the stimulation magnitude until the participant described the sensation as uncomfortable or the maximum safety or hardware limit was reached. For ICMS, pulse amplitude was varied to find the maximum comfortable limit, and the safety limit was 100 µA. For PNS, pulse width was varied to find the maximum comfortable limit, and the hardware limit was 250 µs.

In each experimental session, one electrode was selected for ICMS, one contact was selected for PNS, and the mechanical indenter was positioned in a single area of the hand. In total, across all sessions, two ICMS electrodes and two PNS contacts were chosen, and the mechanical indenter was positioned in four different locations of the hand that spatially overlapped with the evoked percept from one of the selected PNS or ICMS channels. The combination of the ICMS electrode, PNS contact, and mechanical indenter position for each session is shown in **Table 2**. After determining the thresholds and maximum comfortable levels for each modality, we determined three stimulation magnitudes for each modality to be at 30%, 60%, and 90% of the respective sensory dynamic range. Each experimental block then consisted of stimuli varying across the three modalities and three stimulation levels in a randomized order. Trials without any stimulation were also randomly included as catch trials. In each session, the participant completed a total of 4-5 blocks of trials, with 5-15 min breaks in between each block. Each block consisted of 30 trials (3 repetitions x 3 modalities x 3 stimulation magnitudes + 3 catch trials). In 11 PNS trials, 1 mechanical indentation trial, and 1 ICMS trial, the participant had involuntary muscle spasm, and these trials were excluded from the analysis.

**Table 2:** Overview of Experimental Sessions. For each session, the days since the implant, mechanical indenter position, PNS contact, and ICMS contact are shown. The participant was asked to rate the intensity, naturalness, and modality of the evoked sensations in all sessions. In addition, the participant was asked to rate either the location or quality of the evoked sensations in a session. This experiment type is shown in column 2.

| Session | Experiment Type | Days Since Implant | Mechanical Indenter Location | PNS Contact | ICMS Electrode |
| --- | --- | --- | --- | --- | --- |
| 1 | Location | 926 | Ring Fingertip | UC4 | ME59 |
| 2 | Quality | 947 | Ring Fingertip | UC4 | ME59 |
| 3 | Quality | 948 | Ring Fingertip | UC4 | LE10 |
| 4 | Location | 954 | Ring Fingertip | UC4 | LE10 |
| 5 | Quality | 980 | Index Fingertip | UC4 | LE10 |
| 6 | Location | 982 | Index Fingertip | UC4 | LE10 |
| 7 | Quality | 995 | Index Fingertip | MC5 | ME59 |
| 8 | Location | 996 | Index Fingertip | MC5 | ME59 |
| 9 | Quality | 1002 | Index Fingertip | UC4 | ME59 |
| 10 | Location | 1003 | Index Fingertip | UC4 | ME59 |
| 11 | Quality | 1030 | Little Finger Base | UC4 | LE10 |
| 12 | Location | 1031 | Little Finger Base | UC4 | LE10 |
| 13 | Quality | 1038 | Palm Near Index | MC5 | LE10 |
| 14 | Location | 1039 | Palm Near Index | MC5 | LE10 |

### Perceptual Measures

After each trial, the participant reported the perceived modality (i.e., which modality of stimulation the participant believed was applied: ICMS, PNS, mechanical indentation, or none), perceived intensity, and perceived naturalness. Both the experimenter and the participant were blinded to the modality being stimulated in each trial. In half of the sessions, the participant was also asked to report the perceived location of the sensation, and in the other half, the participant was also asked to report the perceived quality of the evoked sensation. Perceived location and perceived quality were never asked in the same session (**Table 2**) due to time constraints.

The participant rated the perceived intensity and the perceived naturalness on self-selected, open-ended scales (**Figure 1**)^92^. For perceived intensity, the participant was instructed to give a rating of zero if no sensation was felt, and to rate a stimulus that felt twice as strong as another stimulus with a number that was twice as large^92^. For perceived naturalness, the participant was instructed to rate the perceived naturalness on a consistent, self-selected scale where 0 is completely unnatural and the maximum number is as natural as touch applied to their face using a dowel. The face was selected as the reference since it was not impaired by the spinal cord injury and thus represents completely natural touch. Note that the participant selected a rating of 10 to correspond with the naturalness of sensation on their face.

The participant verbally reported the perceived location of the evoked projected fields from ICMS, PNS, or mechanical indentation input based on a computer-presented image of the hand divided into sections^15,20^ **(Figure 1, Supplementary Figure 2**). Any number of location regions could be selected. The participant reported the perceived quality of the evoked sensations by verbally indicating which set of words from a provided word bank best described the type of sensation felt^12^ (**Figure 1**). Our word bank was designed to include sensation descriptors reported in prior ICMS and PNS studies^12,15,25–27,93^, as well as to represent all classes of somatosensation, including pain, touch, proprioception, and temperature^38,94^. We also added a few descriptors to the word bank at the request of the participant, including ‘gentle’ and ‘sudden’. The participant was instructed to select as many words as needed to describe the sensation and to use a consistent definition for each word throughout the experiment^12^. The participant was interviewed about their definitions for each descriptor word, and these definitions are presented in **Supplementary Table 2**. To understand the perceived quality of the participant’s sense of natural touch, we applied indentation to the participant’s cheek - a location above the sensory-incomplete SCI impairment. The indentation was applied manually with a dowel at a low, medium, and high depth at least 5 times, and the participant was asked to describe the quality of these sensations using the same word bank.

### Data Analysis

#### Location

Projected fields were plotted across a session as a percentage of each pixel that was selected by the participant across all the trials (**Figure 2A**, **Supplementary Figure 2**). Darker shades indicate the pixel wasselected in more trials. The projected field area was calculated as the percentage of the total hand area (%), both ventral and dorsal, by dividing the number of pixels in the regions reported for each stimulus by the total number of pixels composing the hand in the presented diagram (**Supplementary Figure 8**). The pooled data across all sessions was then compared across the stimulation modalities (**Figure 2C**). To observe the effect of stimulation magnitude for each stimulation modality, separate analyses were done for each contact/electrode/location (**Supplementary Figure 3**). We also investigated if the projected field area would differ when the percept locations evoked by each stimulation modality were spatially disparate vs when the percepts evoked by ICMS or PNS overlapped with those produced by mechanical indentation of the hand. Only one stimulation modality was delivered in each trial, and the term ‘Overlap’ in **Figure 2D** refers to the spatial overlap in evoked percept location within a session. Comparison of “overlapping” and spatial disparate conditions was done separately for each modality and each contact/electrode.

#### Perceived Intensity

All the perceived intensity ratings were normalized by the average intensity rating in each block. The ratings were then pooled across sessions to compare between the modalities (**Figure 3A**) and across the three stimulation magnitudes for each modality (**Figure 3B**). We also calculated the number of trials where the participant reported no sensation as a percentage of the total number of trials across sessions per modality (**Figure 3C**). The standard deviation of the reported intensities was calculated for each session and stimulation level to quantify the consistency of the perceived intensity ratings for each stimulation modality (**Figure 3D**, **Supplementary Figure 6**).

#### Quality

For each session, we calculated the percentage of trials in which each descriptor was chosen for a given stimulation modality and stimulation magnitude condition. We then plotted the average for each stimulation modality across stimulation magnitudes in **Figure 4A**. We also wanted to observe the effect of stimulation magnitude on perceived quality in each stimulation modality. Towards that goal, we selected four descriptors (‘gentle’, ‘startling’, ‘movement (joint)’ and ‘stinging’) that had greater than 5% difference in the percentage of trials reported across the stimulation magnitude levels for any modality **(Figure 4B).** To calculate the quality dissimilarities between pairs of stimulation conditions, we first removed redundant or highly correlated descriptors within each session’s dataset. For each session, we performed principal component analysis on the quality percentages across all trials in the session and selected the first 4 principal components, which explained > 99% of the variance^12^. Then, the quality percentages for the session were projected onto the first 4 principal components, and the pairwise Euclidian distances of the PCA projections between the stimulation conditions were calculated between every combination of stimulation modality and magnitude. So, for each session, we calculated a total of nine distance values (3 modality pairs x 3 magnitudes).^12^. This process was then repeated for each session in which quality was reported, resulting in a total of 7 distance matrices (see **Table 2**). Distances for specific pairs of conditions (PNS-ICMS, ICMS-Mech, and PNS-Mech) were then pooled across sessions (**Figure 4C**)^12^.

#### Naturalness

The naturalness ratings were pooled across sessions to compare across the stimulation modalities (**Figure 5A**) and across the stimulation magnitudes for each modality (**Figure 5B**).

#### Modality Detection

To evaluate the participant’s accuracy of identifying the stimulation modality, we calculated the percentage of trials where the perceived modality matched the stimulated modality, pooled across all sessions. We also calculated the confusion matrix to determine the accuracy of each modality separately and the percentage of trials where one modality was incorrectly identified as a different modality or no sensation (**Supplementary Figure 5**).

#### Correlation of Perceived Intensity and Quality with Naturalness

We performed linear regression to determine the correlation of perceived intensity with perceived naturalness across all trials (all three stimulation magnitudes and all three modalities) (**Figure 6**Error! Reference source not found.**A**). Because we observed that stimulation magnitude correlated with the perceived naturalness, we then performed a stepwise regression, to determine whether perceived intensity had any additional effect on perceived naturalness after the effect of stimulation magnitude was regressed out. To determine the correlation between perceived quality and perceived naturalness, we performed a multiple linear regression to predict the average perceived naturalness of each stimulation condition (the combination of modality and stimulation magnitude) for each session based on the weighted combination of the quality principal component projections for that condition and session. Because there were four principal components to describe the quality data, and thus four terms in the multiple linear regression, we plotted the predicted naturalness based on this regression model on the x-axis and the actual reported perceived naturalness on the y-axis in **Figure 6B** to demonstrate the relationship between these perceptual variables.

#### Statistical Analysis

One-way ANOVAs with Tukey HSD post-hoc comparisons were used to compare the perceived intensity, location area, quality dissimilarity, and naturalness ratings across the three stimulation modalities and across the three stimulation magnitudes within a modality. For naturalness, we also performed a one-sample Wilcoxon signed-rank test to compare the naturalness ratings for each modality to the natural touch rating for the face (reported as 10) (**Figure 5A**). For comparing the effect of spatial overlap of projected fields on evoked location area, perceived intensity and naturalness, we performed the Wilcoxon rank-sum test. The default statistical significance level for these statistical tests was p = 0.05.

#### Partial Least Square Structural Equation Modeling

Partial least squares structural equation models^66,67^ (PLS-SEM) were fit to the perceptual data to examine the underlying structure of the relationships among stimulation and perceptual variables. All data collected across all experimental sessions was pooled for this analysis. We fit three different structural equation models for each stimulation modality. Model A consisted of direct effects of stimulation magnitude on each of the perceptual variables: perceived intensity, naturalness, and quality. Model B hypothesized that stimulation magnitude had direct effects on perceived intensity and perceived quality, and indirect effects on perceived naturalness, which were mediated by perceived intensity and quality. Model C combined Models A and B, resulting in both direct and indirect effects of stimulation magnitude on perceived naturalness, as well as direct effects of stimulation magnitude on perceived intensity and quality. Perceived quality was first converted to the principal component space, as described before, by projecting the descriptor report frequencies onto the first four principal components (which described >99% of the variance) for each session separately. These four principal components then served as four indicator variables which combined to measure the construct of perceived quality in the PLS-SEM. The other perceptual variables (perceived quality and intensity) were assumed to be directly measured by a single metric: their numerical rating on an open-ended scale normalized to the block mean. The models were fit using the SmartPLS software^95^. The output metrics were the standardized root mean square residuals (SRMR), the R^2^ values for naturalness, and the model connection weights^96^.

## List of Supplementary Materials

Supplementary Figure 1-8

Supplementary Table 1-3

## Data Availability

All data produced in the present study are available upon reasonable request to the authors

## Acknowledgements

The authors would like to acknowledge the participant and his family for their time, commitment, and intellectual contributions to this study. The authors would like to acknowledge the Cleveland FES Center for medical illustration and administrative support. The authors thank Charles Greenspon for reading an earlier version of the manuscript and providing valuable feedback. The authors would also like to acknowledge John Krall and Eric Chen for helping design the hardware infrastructure for the mechanical indentation device.

## Author Contribution

E.L.G., B.C.H., B.J.S., and P.B. designed the experiments. B.C.H. and P.B. coded the experiments. B.C.H., B.J.S. and P.B. performed data collection. B.C.H., R.B., and B.J.S. analyzed the data and generated figures. B.C.H., R.B., B.J.S., P.B., and E.L.G. wrote the manuscript. A.K.O and W.D.M. provided engineering and clinical support. J.P.M., J.S., and E.H. performed surgical procedures and managed medical care. A.B.A., J.P.M., and R.F.K. conceived of and oversaw the clinical trial. E.L.G. and A.B.A. interpreted the results and provided scientific oversight of the study. All authors contributed to manuscript revision. E.L.G. supervised the study.

## One Sentence Summary

Cortical stimulation is a better modality for conveying naturalistic touch and peripheral nerve stimulation is more reliable than cortical stimulation.

## References

1. Saudabayev, A., Rysbek, Z., Khassenova, R. & Atakan Varol, H. Human grasping database for activities of daily living with depth, color and kinematic data streams. Scientific Data 2018 5:1 5, 1–13 (2018).

2. Johansson, R. S. & Flanagan, J. R. Coding and use of tactile signals from the fingertips in object manipulation tasks. Nature Reviews Neuroscience 2009 10:5 10, 345–359 (2009).

3. Augurelle, A. S., Smith, A. M., Lejeune, T. & Thonnard, J. L. Importance of cutaneous feedback in maintaining a secure grip during manipulation of hand-held objects. J Neurophysiol 89, 665–671 (2003).

4. Klatzky, R. L. & Lederman, S. J. Stages of manual exploration in haptic object identification. Percept Psychophys 52, 661–670 (1992).

5. Crossman, M. W. Sensory deprivation in spinal cord injury - an essay. Spinal Cord 1996 34:10 34, 573–577 (1996).

6. Lenggenhager, B., Pazzaglia, M., Scivoletto, G., Molinari, M. & Aglioti, S. M. The Sense of the Body in Individuals with Spinal Cord Injury. PLoS One 7, e50757 (2012).

7. Graczyk, E. L. & Tyler, D. J. Somatosensory Neuromodulation with a Focus Towards Clinical Systems. Handbook of Neuroengineering 1–55 (2022) doi:10.1007/978-981-15-2848-4_92-1.

8. Fernández, E. et al. Acute human brain responses to intracortical microelectrode arrays: Challenges and future prospects. Front Neuroeng 7, 92611 (2014).

9. Klein, E., Brown, T., Sample, M., Truitt, A. R. & Goering, S. Engineering the Brain: Ethical Issues and the Introduction of Neural Devices. Hastings Center Report 45, 26–35 (2015).

10. Prodanov, D. & Delbeke, J. Mechanical and biological interactions of implants with the brain and their impact on implant design. Front Neurosci 10, 174893 (2016).

11. Tan, D. W. et al. A neural interface provides long-term stable natural touch perception. Sci Transl Med 6, (2014).

12. Graczyk, E. L., Christie, B. P., He, Q., Tyler, D. J. & Bensmaia, S. J. Frequency Shapes the Quality of Tactile Percepts Evoked through Electrical Stimulation of the Nerves. Journal of Neuroscience 42, 2052–2064 (2022).

13. Graczyk, E. L., Resnik, L., Schiefer, M. A., Schmitt, M. S. & Tyler, D. J. Home Use of a Neural-connected Sensory Prosthesis Provides the Functional and Psychosocial Experience of Having a Hand Again. Scientific Reports 2018 8:1 8, 1–17 (2018).

14. Schiefer, M. A., Graczyk, E. L., Sidik, S. M., Tan, D. W. & Tyler, D. J. Artificial tactile and proprioceptive feedback improves performance and confidence on object identification tasks. PLoS One 13, e0207659 (2018).

15. Flesher, S. N. et al. Intracortical microstimulation of human somatosensory cortex. Sci Transl Med 8, (2016).

16. Greenspon, C. M. et al. Biomimetic multi-channel microstimulation of somatosensory cortex conveys high resolution force feedback for bionic hands. bioRxiv 2023.02.18.528972 (2023) doi:10.1101/2023.02.18.528972.

17. Hobbs, T. G. et al. Biomimetic stimulation patterns drive natural artificial touch percepts using intracortical microstimulation in humans. medRxiv 2024.07.31.24311276 (2024) doi:10.1101/2024.07.31.24311276.

18. Christie, B. et al. Perceived timing of cutaneous vibration and intracortical microstimulation of human somatosensory cortex. Brain Stimul 15, 881–888 (2022).

19. Raspopovic, S., Valle, G. & Petrini, F. M. Sensory feedback for limb prostheses in amputees. Nature Materials 2021 20:7 20, 925–939 (2021).

20. Salas, M. A. et al. Proprioceptive and cutaneous sensations in humans elicited by intracortical microstimulation. Elife 7, (2018).

21. Fisher, L. E., Gaunt, R. A. & Huang, H. Sensory restoration for improved motor control of prostheses. Curr Opin Biomed Eng 28, 100498 (2023).

22. Quick, K. M., Weiss, J. M., Clemente, F., Gaunt, R. A. & Collinger, J. L. Intracortical Microstimulation Feedback Improves Grasp Force Accuracy in a Human Using a Brain-Computer Interface. Proceedings of the Annual International Conference of the IEEE Engineering in Medicine and Biology Society, EMBS 2020-July, 3355–3358 (2020).

23. Mastinu, E. et al. Neural feedback strategies to improve grasping coordination in neuromusculoskeletal prostheses. Scientific Reports 2020 10:1 10, 1–14 (2020).

24. Clemente, F. et al. Intraneural sensory feedback restores grip force control and motor coordination while using a prosthetic hand. J Neural Eng 16, 026034 (2019).

25. Su, S. et al. Sensory feedback by peripheral nerve stimulation improves task performance in individuals with upper limb loss using a myoelectric prosthesis. J Neural Eng 13, 016001 (2015).

26. Hughes, C. L. et al. Perception of microstimulation frequency in human somatosensory cortex. Elife 10, (2021).

27. Valle, G. et al. Tactile edges and motion via patterned microstimulation of the human somatosensory cortex. Science (1979) 387, 315–322 (2025).

28. Valle, G. et al. Comparison of linear frequency and amplitude modulation for intraneural sensory feedback in bidirectional hand prostheses. Scientific Reports 2018 8:1 8, 1–13 (2018).

29. Valle, G. et al. Sensitivity to temporal parameters of intraneural tactile sensory feedback. J Neuroeng Rehabil 17, 1–12 (2020).

30. Flesher, S. N. et al. A brain-computer interface that evokes tactile sensations improves robotic arm control. Science (1979) 372, 831–836 (2021).

31. Raspopovic, S. et al. Bioengineering: Restoring natural sensory feedback in real-time bidirectional hand prostheses. Sci Transl Med 6, (2014).

32. D’Anna, E. et al. A closed-loop hand prosthesis with simultaneous intraneural tactile and position feedback. Sci Robot 4, (2019).

33. Bensmaia, S. J. & Hollins, M. The vibrations of texture. Somatosens Mot Res 20, 33–43 (2003).

34. Skedung, L. et al. Feeling Small: Exploring the Tactile Perception Limits. Scientific Reports 2013 3:1 3, 1–6 (2013).

35. Charkhkar, H. et al. Stability and selectivity of a chronic, multi-contact cuff electrode for sensory stimulation in human amputees. J Neural Eng 12, 026002 (2015).

36. Herring, E. Z. et al. Reconnecting the Hand and Arm to the Brain: Efficacy of Neural Interfaces for Sensorimotor Restoration After Tetraplegia. Neurosurgery 94, 864–874 (2024).

37. Graczyk, E. L. et al. The neural basis of perceived intensity in natural and artificial touch. Sci Transl Med 8, (2016).

38. Guest, S. et al. The development and validation of sensory and emotional scales of touch perception. Atten Percept Psychophys 73, 531–550 (2011).

39. Valle, G. et al. Biomimetic computer-to-brain communication enhancing naturalistic touch sensations via peripheral nerve stimulation. Nature Communications 2024 15:1 15, 1–1 (2024).

40. Petrini, F. M. et al. Six-Month Assessment of a Hand Prosthesis with Intraneural Tactile Feedback. Ann Neurol 85, 137–154 (2019).

41. Valle, G. et al. Biomimetic Intraneural Sensory Feedback Enhances Sensation Naturalness, Tactile Sensitivity, and Manual Dexterity in a Bidirectional Prosthesis. Neuron 100, 37–45.e7 (2018).

42. Gardner, E. P. & Martin, J. H. Coding of Sensory Information. vol. 4 (2000).

43. Ajiboye, A. B. et al. Restoration of reaching and grasping movements through brain-controlled muscle stimulation in a person with tetraplegia: a proof-of-concept demonstration. The Lancet 389, 1821–1830 (2017).

44. Finnerup, N. B., Johannesen, I. L., Fuglsang-Frederiksen, A., Bach, F. W. & Jensen, T. S. Sensory function in spinal cord injury patients with and without central pain. Brain 126, 57–70 (2003).

45. Ikegaya, N. et al. A novel robot-assisted method for implanting intracortical sensorimotor devices for brain-computer interface studies: principles, surgical techniques, and challenges. J Neurosurg 1, 1–9 (2024).

46. Jo, H. J. et al. Multisite Hebbian Plasticity Restores Function in Humans with Spinal Cord Injury. Ann Neurol 93, 1198–1213 (2023).

47. Jo, H. J. & Perez, M. A. Corticospinal-motor neuronal plasticity promotes exercise-mediated recovery in humans with spinal cord injury. Brain 143, 1368–1382 (2020).

48. Grover, F. M., Chen, B. & Perez, M. A. Increased paired stimuli enhance corticospinal-motoneuronal plasticity in humans with spinal cord injury. J Neurophysiol 129, 1414–1422 (2023).

49. Hu, L. T. & Bentler, P. M. Cutoff criteria for fit indexes in covariance structure analysis: Conventional criteria versus new alternatives. Struct Equ Modeling 6, 1–55 (1999).

50. Hughes, C. L., Flesher, S. N. & Gaunt, R. A. Effects of stimulus pulse rate on somatosensory adaptation in the human cortex. Brain Stimul 15, 987–995 (2022).

51. Devivo, M. J. Epidemiology of traumatic spinal cord injury: trends and future implications. Spinal Cord 2012 50:5 50, 365–372 (2012).

52. Abraira, V. E. & Ginty, D. D. The Sensory Neurons of Touch. Neuron 79, 618–639 (2013).

53. Delhaye, B. P., Long, K. H. & Bensmaia, S. J. Neural Basis of Touch and Proprioception in Primate Cortex. Compr Physiol 8, 1575–1602 (2018).

54. Pei, Y. C., Denchev, P. V., Hsiao, S. S., Craig, J. C. & Bensmaia, S. J. Convergence of submodality-specific input onto neurons in primary somatosensory cortex. J Neurophysiol 102, 1843–1853 (2009).

55. Suresh, A. K. et al. Sensory computations in the cuneate nucleus of macaques. Proc Natl Acad Sci U S A 118, e2115772118 (2021).

56. Schiefer, M. A., Triolo, R. J. & Tyler, D. J. A model of selective activation of the femoral nerve with a flat interface nerve electrode for a lower extremity neuroprosthesis. IEEE Transactions on Neural Systems and Rehabilitation Engineering 16, 195–204 (2008).

57. Schiefer, M. A., Tyler, D. J. & Triolo, R. J. Probabilistic modeling of selective stimulation of the human sciatic nerve with a flat interface nerve electrode. Proceedings of the Annual International Conference of the IEEE Engineering in Medicine and Biology Society, EMBS 4068–4071 (2011) doi:10.1109/IEMBS.2011.6091011.

58. Okorokova, E. V., He, Q. & Bensmaia, S. J. Biomimetic encoding model for restoring touch in bionic hands through a nerve interface. J Neural Eng 15, 066033 (2018).

59. Saal, H. P., Delhaye, B. P., Rayhaun, B. C. & Bensmaia, S. J. Simulating tactile signals from the whole hand with millisecond precision. Proc Natl Acad Sci U S A 114, E5693–E5702 (2017).

60. Jiang, L. et al. A biomimetic electrical stimulation strategy to induce asynchronous stochastic neural activity. J Neural Eng 17, 046019 (2020).

61. Greenspon, C. M. et al. Evoking stable and precise tactile sensations via multi-electrode intracortical microstimulation of the somatosensory cortex. Nature Biomedical Engineering 2024 1–17 (2024) doi:10.1038/s41551-024-01299-z.

62. Fifer, M. S. et al. Intracortical Somatosensory Stimulation to Elicit Fingertip Sensations in an Individual with Spinal Cord Injury. Neurology 98, E679–E687 (2022).

63. Clark, G. A. et al. Using multiple high-count electrode arrays in human median and ulnar nerves to restore sensorimotor function after previous transradial amputation of the hand. Annu Int Conf IEEE Eng Med Biol Soc 2014, 1977–1980 (2014).

64. Overstreet, C. K., Klein, J. D. & Helms Tillery, S. I. Computational modeling of direct neuronal recruitment during intracortical microstimulation in somatosensory cortex. J Neural Eng 10, 066016 (2013).

65. Stoney, S. D., Thompson, W. D. & Asanuma, H. Excitation of pyramidal tract cells by intracortical microstimulation: effective extent of stimulating current. 10.1152/jn.1968.31.5.659 31, 659–669 (1968).

66. Kumaravelu, K., Sombeck, J., Miller, L. E., Bensmaia, S. J. & Grill, W. M. Stoney vs. Histed: Quantifying the spatial effects of intracortical microstimulation. Brain Stimul 15, 141–151 (2022).

67. Histed, M. H., Bonin, V. & Reid, R. C. Direct Activation of Sparse, Distributed Populations of Cortical Neurons by Electrical Microstimulation. Neuron 63, 508–522 (2009).

68. Martuzzi, R., van der Zwaag, W., Farthouat, J., Gruetter, R. & Blanke, O. Human finger somatotopy in areas 3b, 1, and 2: A 7T fMRI study using a natural stimulus. Hum Brain Mapp 35, 213–226 (2014).

69. Stewart, J. D. Peripheral nerve fascicles: Anatomy and clinical relevance. Muscle Nerve 28, 525–541 (2003).

70. Pleger, B. & Villringer, A. The human somatosensory system: From perception to decision making. Prog Neurobiol 103, 76–97 (2013).

71. Graczyk, E. L., Delhaye, B. P., Schiefer, M. A., Bensmaia, S. J. & Tyler, D. J. Sensory adaptation to electrical stimulation of the somatosensory nerves. J Neural Eng 15, 046002 (2018).

72. McGlone, F., Wessberg, J. & Olausson, H. Discriminative and Affective Touch: Sensing and Feeling. Neuron 82, 737–755 (2014).

73. Murphy, S. & Dalton, P. Out of Touch? Visual Load Induces Inattentional Numbness. J Exp Psychol Hum Percept Perform 42, 761–765 (2016).

74. Mun, S., Whang, M., Park, S. & Park, M. C. Effects of mental workload on involuntary attention: A somatosensory ERP study. Neuropsychologia 106, 7–20 (2017).

75. Christie, B. P. et al. Visual inputs and postural manipulations affect the location of somatosensory percepts elicited by electrical stimulation. Scientific Reports 2019 9:1 9, 1–14 (2019).

76. Donati, E. & Valle, G. Neuromorphic hardware for somatosensory neuroprostheses. Nature Communications 2024 15:1 15, 1–18 (2024).

77. Gonzalez, M., Bismuth, A., Lee, C., Chestek, C. A. & Gates, D. H. Artificial referred sensation in upper and lower limb prosthesis users: a systematic review. J Neural Eng 19, 051001 (2022).

78. Bensmaia, S. J. Biological and bionic hands: natural neural coding and artificial perception. Philosophical Transactions of the Royal Society B: Biological Sciences 370, (2015).

79. Bjånes, D. A. & Moritz, C. T. Design of intracortical microstimulation patterns to control the location, intensity, and quality of evoked sensations in human and animal models. Somatosensory Feedback for Neuroprosthetics 479–506 (2021) doi:10.1016/B978-0-12-822828-9.00018-6.

80. Bjånes, D. A. et al. Multi-channel intra-cortical micro-stimulation yields quick reaction times and evokes natural somatosensations in a human participant. medRxiv 2022.08.08.22278389 (2022) doi:10.1101/2022.08.08.22278389.

81. Bell-Krotoski, J. A., Fess, E. E., Figarola, J. H. & Hiltz, D. Threshold Detection and Semmes-Weinstein Monofilaments. Journal of Hand Therapy 8, 155–162 (1995).

82. Nolan, M. F. Two-Point Discrimination Assessment in the Upper Limb in Young Adult Men and Women. Phys Ther 62, 965–969 (1982).

83. Charkhkar, H. et al. The design of and chronic tissue response to a composite nerve electrode with patterned stiffness. J Neural Eng 14, 036022 (2017).

84. McCreery, D., Han, M., Pikov, V. & Miller, C. Configuring intracortical microelectrode arrays and stimulus parameters to minimize neuron loss during prolonged intracortical electrical stimulation. Brain Stimul 14, 1553–1562 (2021).

85. McCreery, D. B., Agnew, W. F., Yuen, T. G. H. & Bullara, L. Charge density and charge per phase as cofactors in neural injury induced by electrical stimulation. IEEE Trans Biomed Eng 37, 996– 1001 (1990).

86. Shannon, R. V. A Model of Safe Levels for Electrical Stimulation. IEEE Trans Biomed Eng 39, 424–426 (1992).

87. Callier, T., Suresh, A. K. & Bensmaia, S. J. Neural Coding of Contact Events in Somatosensory Cortex. Cerebral Cortex 29, 4613–4627 (2019).

88. Cohen, R. H. & Vierck, C. J. Relationships between touch sensations and estimated population responses of peripheral afferent mechanoreceptors. Exp Brain Res 94, 120–130 (1993).

89. Tabot, G. A. et al. Restoring the sense of touch with a prosthetic hand through a brain interface. Proc Natl Acad Sci U S A 110, 18279–18284 (2013).

90. Poulos, D. A. et al. The neural signal for the intensity of a tactile stimulus. Journal of Neuroscience 4, 2016–2024 (1984).

91. Burgess, P. R. et al. The neural signal for skin indentation depth. I. Changing indentations. Journal of Neuroscience 3, 1572–1585 (1983).

92. Jones, L. A. & Tan, H. Z. Application of psychophysical techniques to haptic research. IEEE Trans Haptics 6, 268–284 (2013).

93. Charkhkar, H. et al. High-density peripheral nerve cuffs restore natural sensation to individuals with lower-limb amputations. J Neural Eng 15, 056002 (2018).

94. Kim, L. H., McLeod, R. S. & Kiss, Z. H. T. A new psychometric questionnaire for reporting of somatosensory percepts. J Neural Eng 15, 013002 (2018).

95. Ringle, C. M., Wende, S., & Becker, J. M. (2024). SmartPLS 4. SmartPLS. - References - Scientific Research Publishing. https://www.scirp.org/reference/referencespapers?referenceid=3813136.

96. Sarstedt, M., Ringle, C. M. & Hair, J. F. Partial Least Squares Structural Equation Modeling. Handbook of Market Research 587–632 (2021) doi:10.1007/978-3-319-57413-4_15/TABLES/8.

