## Supplemental for "Perceptual Differences between Cortical and Peripheral Stimulation Strategies for Sensory Restoration"

### Supplemental Materials

**Supplementary Table 1: Performance on clinical tests of sensory function after implantation of the ReHAB system.** The Semmes-Weinstein monofilament test and two-point discrimination tests were conducted on each finger and palm of the right hand. For both tests, higher values indicate worse performance. The first column denotes typical normal values in the healthy population[1,2].

| | Normal | Thumb | Index | Middle | Ring | Little | Palm center | Ring palm crease |
|---|---|---|---|---|---|---|---|---|
| **Monofilament Test (mm)** | 2.83 | 4.31 | 4.17 | 4.17 | 4.17 | 4.17 | 4.56 | 4.31 |
| **Two-point Discrimination Test (mm)** | 2.57 | 12 | 7 | 7 | 5 | 5 | N/A | N/A |

**Supplementary Table 2: Quality Descriptor Definitions.** The participant was interviewed about his definitions for each quality descriptor word in the provided word bank, as well as for the words the participant requested to be added to the word bank, which are denoted by an asterisk. The instructions provided to the participant were: "In your own words, describe what these quality words mean to you." The participant's response was transcribed verbatim and then cleaned to remove fillers. The highlighted rows denote the descriptors that the participant actually reported during the experiment across the three stimulation modalities.

| Quality Word Provided in Word Bank | Participant's Description of Word Definition |
|---|---|
| Touch | "Touch to me is if I perceive anything making contact or simulating making contact with my skin." |
| Pressure | "Pressure would be if I feel as if someone is pushing into my cheek… similar sensation in my arm or hand or someone applying pressure into my hand." |
| Tingling | "Tingling, in my opinion, I use it mostly to describe [what it feels like] after being in contact or someone applying pressure or touch. I feel like I get tingling in that area, not for long but for a little bit, almost to let me know in my brain where they applied stimulation to. Greater pressure results in much more residual tingling, and so, that's I think how I use tingling - as a way to describe that." |
| Squeezing | "Squeezing: I use that to describe the sensation of… having the feeling as if someone is squeezing both sides of my fingertip at the same time. And so when I use that – squeezing - I am referencing that. It is a whole finger sensation, in that I am not able to differentiate between whether it is the palm side or that top side, being that both sides are applied contact at the same time." |

| | |
|---|---|
| Buzzing | “With buzzing, it is kind of that sensation or feeling of the FES (functional electrical stimulation) going through my hand or arm depending on where it is. I always call it current-y. …Buzzing almost has that sensation of like it's not still, it is very much moving - like vibrating kind of… similar in the way I am interpreting the FES sensation.” |
| Electrical | “Electrical feels much more like a flow - I guess is what I think of as like the current flowing up and down. It seems like it is flowing forwards and back and back and forward in the area that we are producing it in.” |
| Movement (joint) | “Movement (joint) is usually what I would classify as me feeling a particular part of my hand or arm kind of shifted from its rested or preset position. So, if I feel that it was in position A, then moved to position B, and maybe goes back to A, then maybe I would say that I felt some sort of movement of the joint.” |
| Stinging | “Stinging would only apply to certain FES (peripheral nerve stimulation) ones (stimuli), when they get high. It's not so much that it is painful or really uncomfortable, more so just like uh, think of like a light stinging. I guess it is in that realm of electrical but maybe now with more of a shocking or stinging type sensation. Now it is actually taking on something that could maybe lead into uncomfortableness or pain, but it is not quite there yet.” |
| Sudden* | “Sudden: If I use that I am referring to the onset of how I feel the stimulation. Meaning if it ICMS or FES (peripheral nerve stimulation), it seems like it just comes on at its set number (intensity). Maybe if it is a medium or high (intensity) it doesn’t go up to it, it doesn’t have a progression, or I don’t even interpret some sort of quick ramp up. It just seems like it's there (at its final intensity) right then out of nowhere.” |
| Gradual* | “For gradual, it's what I use for mechanical indentation because I think, or I'm pretty sure I know, my intact sensation being more robust, gives me more feedback or gives me more complexity in the sensation so I can actually feel the mechanical indentation. Now it’s a very quick ramp up, but it's much more able to be interpreted as a gradual touch… vs feeling it all happen as one sudden event.” |
| Startling* | “Startling: I would classify as probably any of the stimuli that seem like they catch me or my brain off guard. As in, I'm not expecting to be startled. Its unexpected, it doesn’t feel as normal of a stimulation and so I would consider it startling.” |
| Gentle* | “Gentle: I use that a lot of times to explain a sensation that is both light, but I mean almost like comfortable, delicate, gentle in the sense that my mind is the opposite of startled. I mean it's (my mind’s) not thrown off by it (the stimulation). It's actually something that is very relaxing. I don’t know about relaxing but very normal or doesn’t seem out of the ordinary or whatnot to feel. It doesn’t pertain to all of them, because I could have a light (intensity) FES (peripheral nerve stimulation) but I would never classify that as gentle. But for the other ones (stimuli), when I feel it, it reminds me if you were to, I guess I would interpret it as a gentle touch on the cheek.” |

| | |
|---|---|
| Uncomfortable | "Uncomfortable would be something that is starting to get into the realm of painful… Something that I usually wouldn't get often, but if I do get a sensation like that, I would say 'yeah, it's starting to get to the realm of what I would classify next as painful.' And most of the times if I'm using it (the word 'uncomfortable'), it's within a spasm that is [happens] after one of the stimulations." |
| Contracting | "Contracting: Really the only time I'm referring to that in this context would be if it (the stimulation) causes me to spasm - one of the induced stimulations causes me to spasm. Which then will contract a lot of my muscles in my arm and sometimes whole body, thus a lot of times leading to an uncomfortable outcome." |
| Painful | "Painful in my opinion would be when we are starting to get to a point where I very much don't want to play around with parameters at that level. And I believe we are far from those standards. Which is why, even when I get a spasm, I don't know, even that I wouldn't call it painful. It's more uncomfortable. But painful again - those parameters are outside of my range. But it would be inducing something consistently at a level that is conflicting harm." |

**Supplementary Table 3: Descriptions of perceived sensations evoked by natural touch applied to the face.** The researcher applied touch stimuli to the left cheek at three depths (low, medium, and high) using a wooden dowel that was approximately 2 mm in diameter. Each depth was presented five times. The participant described the perceived intensity, naturalness, and quality of the evoked sensations using the same methods as described in the main text. The average perceived intensity and naturalness scores, and the quality descriptors reported are shown in the table.

| | **Intensity** | **Naturalness** | **Quality** |
|---|---|---|---|
| **Low depth** | 2 | 10 | Touch, pressure, gradual, gentle |
| **Medium depth** | 4 | 10 | Touch, pressure, gradual |
| **High depth** | 6.25 | 10 | Touch, pressure, gradual |

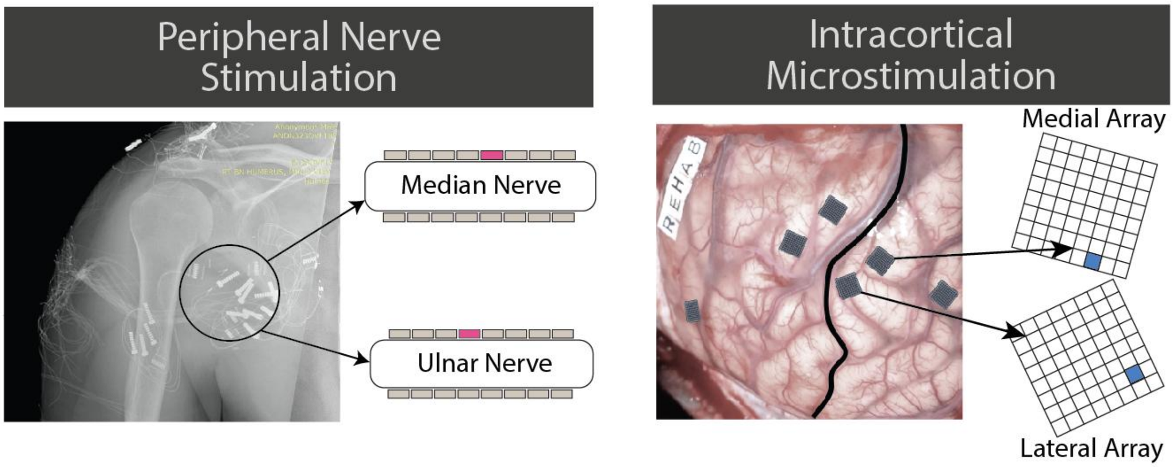


**Supplementary Figure 1:** Placement of the peripheral electrodes for peripheral nerve stimulation (left) and intracortical arrays for intracortical microstimulation (right). The 16-contact composite flat interface nerve electrodes (C-FINEs) were placed around the median and ulnar nerves (shown in post-operative X-ray on left). The fifth median nerve contact and the fourth ulnar nerve contact were selected to deliver stimulation for this study (highlighted in pink). Two 64-channel microelectrode arrays were placed in Brodmann's area 1 of the primary somatosensory cortex (shown on right). The contacts selected to deliver stimulation in this study in the medial and lateral sensory arrays are highlighted in blue.

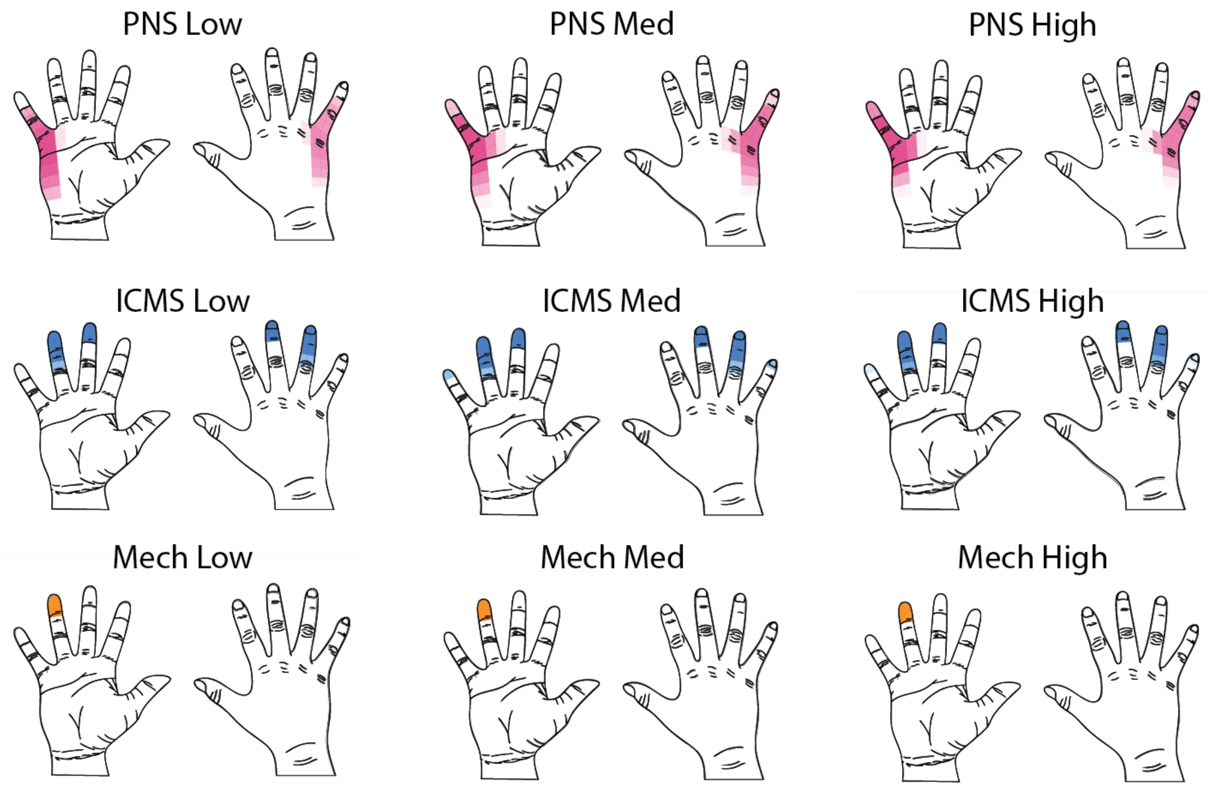


**Supplementary Figure 2:** Examples of projected fields for each stimulation modality across the three stimulation magnitudes (low, medium, and high). The projected fields for PNS (pink) occasionally expanded distally (e.g., toward the little fingertip) and laterally (e.g., toward the base of the ring finger) as stimulation magnitude increased. The projected fields for ICMS (blue) occasionally expanded laterally (e.g., towards the pinky fingertip) as stimulation magnitude increased. The projected fields for mechanical indentation (orange) were always consistent across all stimulation magnitudes.

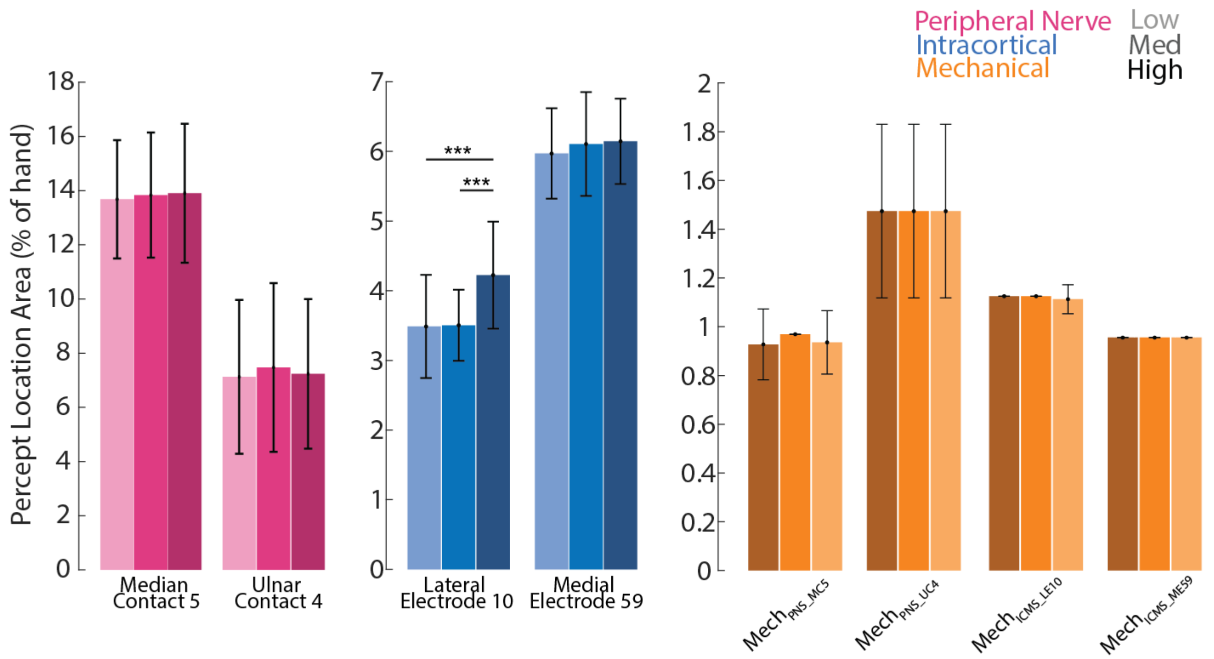


**Supplementary Figure 3:** Impact of stimulation magnitude on the size of the projected fields from the three stimulation modalities. The percept sizes for each contact, electrode, and mechanical indenter position were calculated for the PNS (pink), ICMS (blue), and mechanical indentation (orange) stimuli, respectively. The light, medium, and dark shades of each color indicate low, medium and high stimulation magnitudes, respectively. The *** indicates p<0.001.

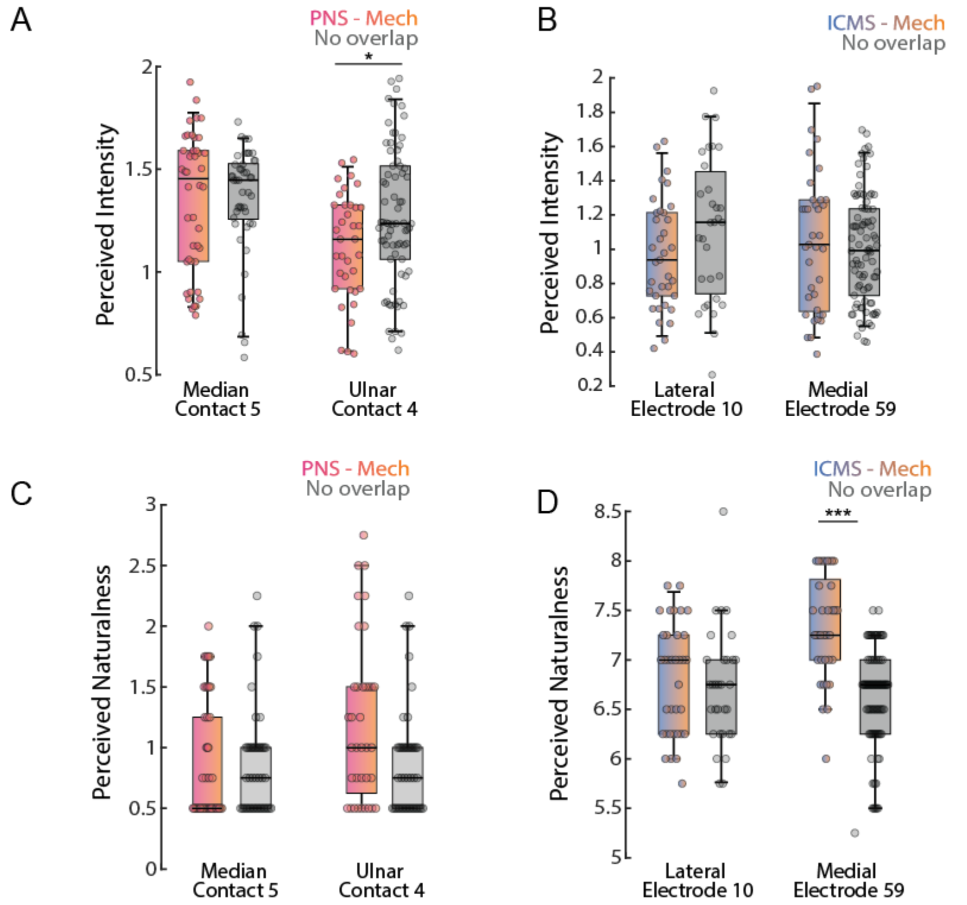


| | | Touch | Pressure | Tingling | Gradual | Sudden | Squeezing | Buzzing | Vibrating | Electrical | Gentle | Startling | Movement (Joint) | Stinging |
|---|---|---|---|---|---|---|---|---|---|---|---|---|---|---|
| Median Contact 5 | PNS-Mech | 0 | 0 | 0 | 0 | 100 | 0 | 100 | 100 | 100 | 0 | 47.22 | 100 | 47.22 |
| | No overlap | 0 | 0 | 95.56 | 0 | 100 | 0 | 100 | 100 | 100 | 0 | 51.11 | 100 | 55.56 |
| Ulnar Contact 4 | PNS-Mech | 0 | 0 | 0 | 0 | 100 | 0 | 100 | 100 | 100 | 0 | 26.67 | 100 | 26.67 |
| | No overlap | 0 | 0 | 0 | 0 | 100 | 0 | 100 | 100 | 100 | 0 | 32.63 | 84.21 | 32.63 |
| Lateral Electrode 10 | ICMS-Mech | 97.78 | 97.78 | 97.78 | 0 | 97.78 | 97.78 | 0 | 0 | 0 | 31.11 | 31.11 | 0 | 0 |
| | No overlap | 97.78 | 97.78 | 97.78 | 0 | 97.78 | 97.78 | 0 | 0 | 0 | 37.78 | 26.67 | 0 | 0 |
| Medial Electrode 59 | ICMS-Mech | 100 | 100 | 100 | 0 | 100 | 100 | 0 | 0 | 0 | 41.67 | 19.44 | 0 | 0 |
| | No overlap | 97.78 | 97.78 | 97.78 | 0 | 97.78 | 97.78 | 0 | 0 | 0 | 33.33 | 21.11 | 0 | 0 |

**Supplementary Figure 4:** Effect of spatial overlap of projected fields across stimulation modalities on perceptual experience. (A) Effect of overlap condition on perceived intensity for PNS. (B) Effect of overlap condition on perceived intensity for ICMS. (C) Effect of overlap condition on perceived naturalness for PNS. (D) Effect of overlap condition on perceived naturalness for ICMS. (E) Effect of overlap condition on perceived quality for PNS. (F) Effect of overlap condition on perceived quality for ICMS. For A, C, and E: Sessions in which the PNS projected field overlapped with the mechanical indenter position (PNS-Mech) were compared to those in which there was no overlap among stimulation conditions (No overlap). For B, D, and F: Sessions in which the ICMS projected field overlapped with the mechanical indenter position (ICMS-Mech) were compared to those in which there was no overlap among stimulation conditions (No overlap). All panels: single asterisk denotes $p<0.05$, triple asterisks denotes $p<0.001$.

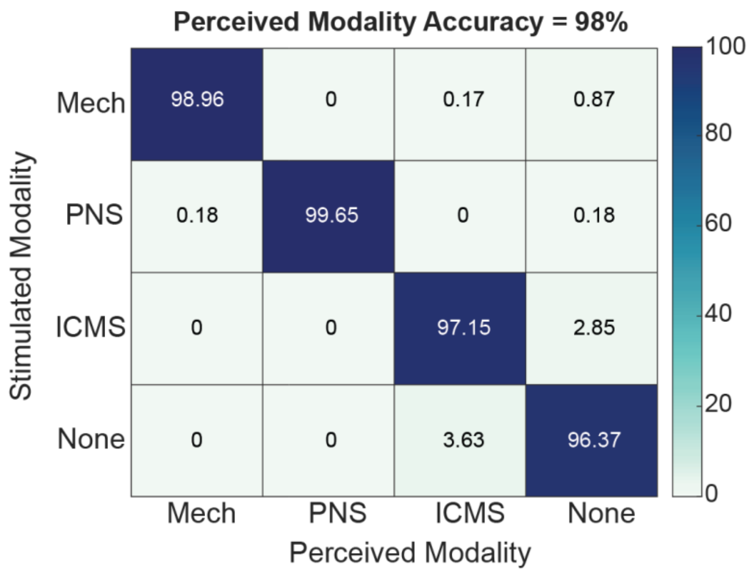


**Supplementary Figure 5:** Accuracy of correctly identifying the stimulation modality across all PNS (n=570), ICMS (n=561), Mech (n=578), and catch trials (n=193). The experimenter and the participant were both blinded to the modality being stimulated in each trial. In this confusion matrix, the horizontal rows indicate the actual modality being stimulated in each trial and the vertical columns indicate the perceived modality reported by the participant. Elements along the diagonal indicate the percentage of trials in which the participant correctly identified the stimulation modality, and the overall accuracy (average of all the diagonal values) is shown in the top of the confusion matrix.

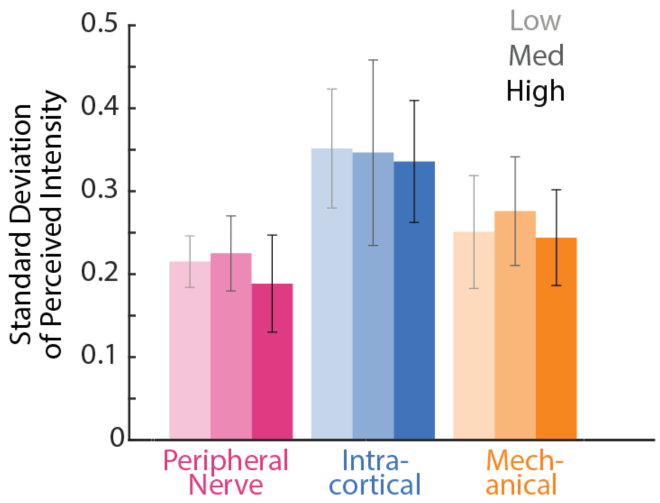


**Supplementary Figure 6:** The standard deviation of the perceived intensities reported in each session for each of the three stimulation magnitudes for each modality. The light to dark shades indicates the low, medium and high stimulation magnitudes, respectively. No statistical significance was present across the stimulation magnitudes for any of the three modalities.

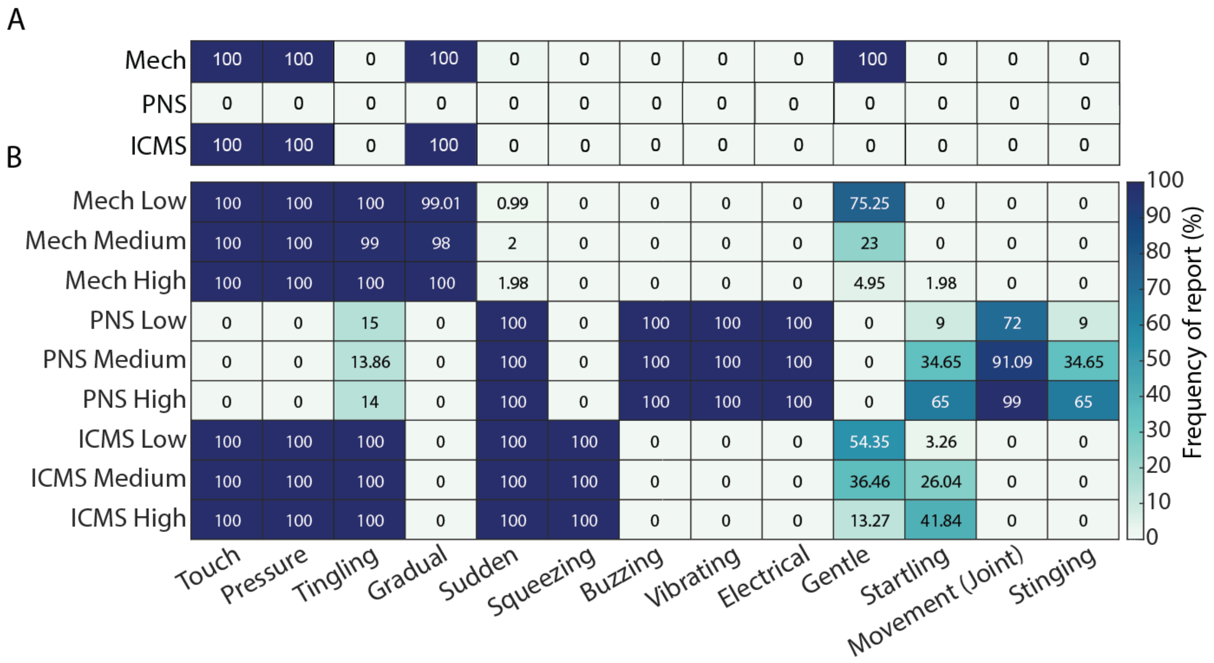


**Supplementary Figure 7:** Effect of stimulation magnitude on the perceived quality of the sensations evoked by each stimulation modality. (A) Frequency each quality descriptor was reported for natural touch applied to the face at low, medium, and high magnitudes (indentation depths). (B) Frequency each quality descriptor was reported across trials for each stimulation magnitude and stimulation modality.

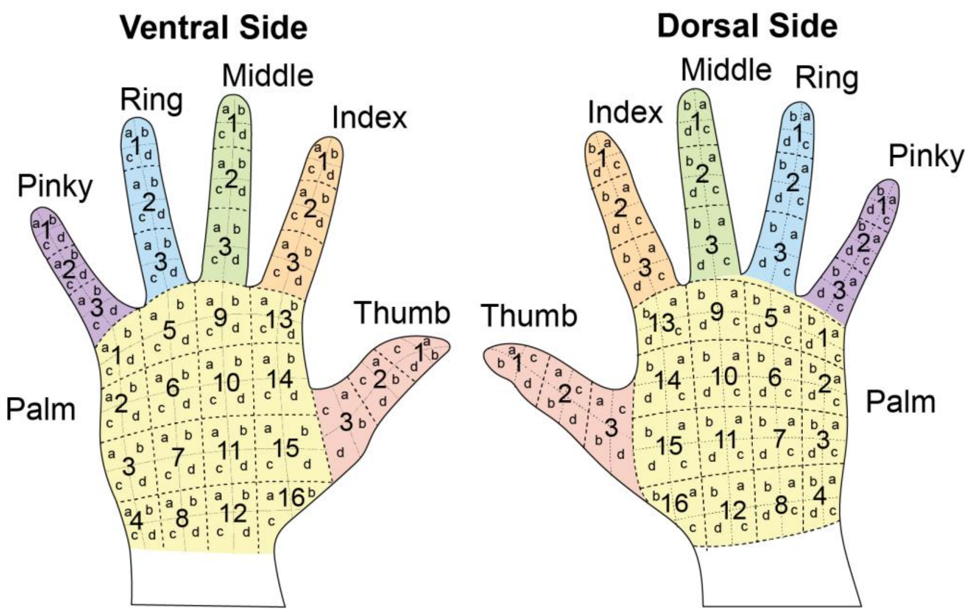


**Supplementary Figure 8:** Discretized hand image used to collect perceived location data. The participant viewed this image on a computer screen after each stimulation trial and verbally reported the numbers and letters for the region(s) he felt the evoked sensation during that trial.
